# Mortality among persons with HIV in the United States during the COVID-19 pandemic: a population-level analysis

**DOI:** 10.1101/2023.04.19.23288817

**Authors:** Alex Viguerie, Ruiguang Song, Karin Bosh, Cynthia M. Lyles, Paul G. Farnham

## Abstract

**Background:** Whether COVID-19 has had a disproportionate impact on mortality among persons with diagnosed HIV (PWDH) in United States is unclear. Through our macro-scale analysis, we seek to better understand how COVID-19 and subsequent behavioral changes affected mortality among PWDH.

**Methods:** We obtained mortality and population size data for the years 2018-2020 from the National HIV Surveillance System (NHSS) for the PWDH population aged ≥13 years in the United States, and from publicly available data for the general population. We computed mortality rates and excess mortality for both the general and PWDH populations. Stratifications by age, race/ethnicity, and sex-at birth were considered. For each group, we determined whether the 2020 mortality rates and mortality risk ratio showed a statistically significant change from 2018-2019.

**Results:** Mortality rates increased in 2020 from 2018-2019 across the general population in all groups. Among PWDH, mortality rates either increased, or showed no statistically significant change. The mortality risk ratio between PWDH and the general population decreased 7.7% in 2020. Approximately 1550 excess deaths occurred among PWDH in 2020, with Black, Hispanic/Latino and PWDH above 55 and older representing the majority of excess deaths.

**Conclusions:** While mortality rates among PWDH increased in 2020 relative to 2018-2019, the increases were smaller than those observed in the general population. This suggests that COVID-19 and resulting behavioral changes among PWDH did not result in disproportionate mortality among PWDH. These findings suggest that COVID-19, and any associated indirect effects, do not represent a proportionally greater risk for PWDH compared to the general population.

## Introduction

The outbreak of COVID-19 in late 2019 has had an unprecedented impact on all facets of life worldwide. Both the disease itself, and its associated disruptions, have led to widespread damage in terms of human well-being. These costs are varied and include serious mental health and economic effects; however, the most immediately apparent effect of COVID-19 has been on human life. In the United States alone, the age-adjusted mortality rate in the general population increased 16.8% in 2020, with life expectancy decreasing a full 1.8 years when compared with 2019 [1], [2]. However, this burden has not been evenly shared, as many groups showed a disproportionately high mortality risk from COVID-19, with the most vulnerable group being people over the age of 65 [1].

Despite many studies on the subject, whether persons with diagnosed HIV (PWDH) are among the groups at higher risk of mortality from COVID-19 remains unclear. Results from meta-analyses and clinical studies have been mixed, with some concluding that PWDH demonstrate an elevated risk of death from COVID-19 compared to the general population [3]–[6] and others finding no such increased risk [7]–[10]. Such studies are difficult to interpret for many reasons, including sample size limitations, differences in the age and biological sex distribution of the PWDH population as compared with the general population, and the fact that mortality is higher among PWDH generally.

It is also important to note that excess mortality due to the COVID-19 pandemic is not solely the result of COVID-19 itself; the direct and indirect effects of lockdowns, as well as other factors perhaps not yet completely understood, led to statistically significant increases in other leading causes of death, including heart disease, diabetes, and unintentional injuries [1], [2]. We may call such deaths *second-order* COVID-19 deaths, as this increased mortality, though not directly caused by COVID-19, is likely related to the systemic disruption caused by the pandemic.

A clear understanding of the larger trends involving COVID-19 related mortality on PWDH is therefore of great importance for future modeling and intervention efforts. Many lines of evidence suggest that COVID-19 and its related second-order effects had significant impacts on the HIV continuum of care [11]–[14]. To design and evaluate the necessary interventions to address these effects, a detailed knowledge of changes in the mortality patterns among PWDH is necessary.

We offer in the present work an analysis based on mortality data among both PWDH and the general population in the United States, both before and during the COVID-19 pandemic. Such an analysis is not inherently affected by the differences in population composition of PWDH and the general population, as such differences may be assumed to be equally present both before and during the COVID-19 pandemic. For example, the PWDH population is over 75% male (assigned at birth), while males comprise 50% of the general population. As males have notably higher mortality rates compared to females in general [15], from this fact alone, we might expect higher mortality among PWDH relative to the general population, even in the absence of COVID-19. Thus, comparing PWDH versus general population mortality in 2020 in isolation may not account for such a difference. However, when comparing differences in PWDH and general population mortality in 2020 *to analogous differences in previous years,* we may assume that the PWDH population having a higher percentage of males affects both the 2020 and previous year data equally, and any observed changes take this into account. Analyzing changes in mortality patterns in both PWDH and the general population groups will allow us to better understand whether COVID-19, as well as the associated behavioral changes, resulted in disproportionate mortality among PWDH compared with the general population. This knowledge is fundamental for understanding both the present and future of HIV in the United States.

## Methods

Data reported to the Center for Disease Control and Prevention’s (CDC) National HIV Surveillance System (NHSS) through December 2021 were used for population and mortality data for PWDH aged ≥13 years [16]. Population and mortality data for the general United States population were taken from the National Center for Health Statistics’ National Vital Statistics System, accessed through the publicly available CDC WONDER database [17].

We also calculated mortality rates across the 13-24, 25-34, 35-44, 45-54, 55-64, and 65+ age groups. Persons under 13 were not considered among PWDH nor in the general population. We also computed mortality rates by assigned sex at birth (male/female) and race/ethnicity, considering White, Hispanic/Latino, and Black or African American (hereafter referred to as Black) populations. We further examined two-way (race/ethnicity and age, and sex-at-birth and age) and three-way (race/ethnicity, sex-at-birth, and age) stratifications of these population groupings.

Mortality rates were calculated as follows: let 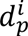 be the number of deaths in a population *p* in the year (or group of years) *i*, and 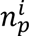 the total population, denoted analogously. Then mortality rates μ for a population *p* a year (or group of years) *i* are given by:

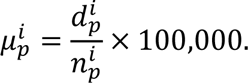

To reduce sensitivity to single-year effects, we consider a combined mortality rate for 2018 and 2019, 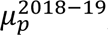, computed as:

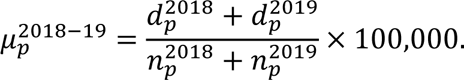

For readability, all reported rates are assumed to be per 100,000 population, even if not explicitly denoted. For all populations *p*, we denote the percent change in mortality rate in 2020 from the 2018-2019 rate as *δ*_*p*_, and compute it as:

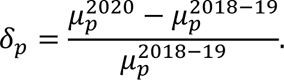

We provide a 95% confidence interval 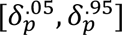 for *δ_p_* under the assumption that the number of deaths in a year follows a binomial distribution, and that the ratio of the 2018-2019 and 2020 mortality rates 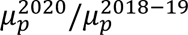 is log-normally distributed.

For each considered group, we then computed the relative mortality risk ratio (RR) between the general population and the PWDH population for each year 2018-19 and 2020 as follows:

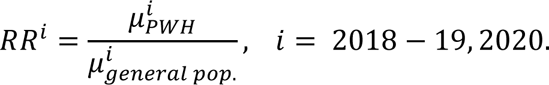

Or, equivalently:

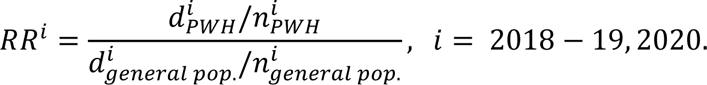

To determine statistical significance, we examined the changes in risk ratio from 2018-2019 (years combined) to 2020. If PWDH showed increased mortality risk due to COVID-19, or its second-order effects, we would expect the RR to be significantly higher in 2020. On the other hand, no change (or decrease) in the risk ratio would imply the opposite. Confidence intervals for the risk-ratio are calculated under the assumption of a log-normal distribution.

To determine whether changes in RR during 2020 were statistically significant, we compute:

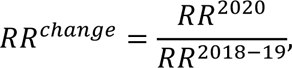

and then determine a p-value *p* by assuming *RR_change_* follows a log-normal distribution, and computing the z-score as:

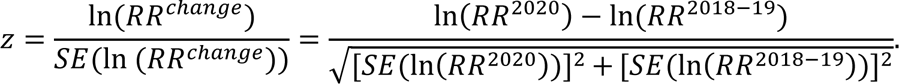

Then *p* is recovered as:

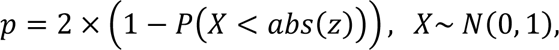

Where *SE* denotes the standard error of the estimated risk ratio, defined here as:

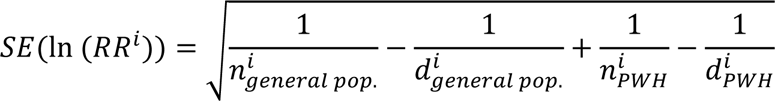

Excess deaths among PWDH during 2020 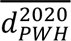were estimated as:

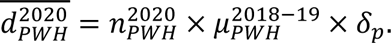

A confidence interval for 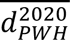was obtained by replacing *δ*_*p*_ in the above expression with the lower- and upper-bounds (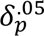 and 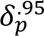, respectively) of the associated confidence interval for *δ*_*p*_. As excess mortality among the general population during 2020 has been studied extensively elsewhere (see [1], [2]), we do not compute this quantity here.

## Results

Among the general population, mortality rates increased notably from 2018-2019 to 2020 across all groups (Table 1). The largest relative increases were among groups aged 54 and under in the general population, due to the lower initial mortality rates (increases of 19.9% or more among the age groups 13-24, 25-34, 35-44, and 45-54 in the general population, compared increases of 17.4% or less among groups aged 55 and older). However, the largest absolute mortality rate increases occurred among groups aged 55 and older. Among PWDH (Table 2), the largest absolute increases in mortality rates occurred in the older age groups. Mortality rates increased from 2018-2019 to 2020 among PWDH for all groups examined except for White females with diagnosed HIV and PWDH ages 13-24 and 45-54, who did not show statistically significant change in mortality rate in 2020.

**Table 1:** Mortality rates^1^ among the general population aged ≥13 years, by age, race/ethnicity, and sex^2^, 2018–2020—United States

| Age | Race/Ethnicity | Sex<sup>2</sup> | Mortality Rate, 2018-19 | Mortality Rate, 2020 | Change from 2018-19 (Absolute) | Change from 2018-19 (%) | Change from 2018-19 (%), 95% confidence interval |
| --- | --- | --- | --- | --- | --- | --- | --- |
| All | All | All | 1,023.6 | 1209.2 | 185.6 | 18.1% | 18.0% — 18.3% |
| All | All | M | 1,077.0 | 1292.6 | 215.6 | 20.0% | 19.8% — 20.2% |
| All | All | F | 972.5 | 1129.3 | 156.9 | 16.1% | 15.9% — 16.4% |
| All | Black/African-American | All | 993.7 | 1288.9 | 295.2 | 29.7% | 29.2% — 30.2% |
| All | Hispanic/Latino | All | 434.1 | 625.6 | 191.5 | 44.1% | 43.4% — 44.8% |
| All | White | All | 1,266.4 | 1443.1 | 176.7 | 14.0% | 13.8% — 14.1% |
| All | Black/African-American | M | 1,097.4 | 1442.8 | 345.4 | 31.5% | 30.8% — 32.2% |
| All | Black/African-American | F | 900.9 | 1151.2 | 250.3 | 27.8% | 27.1% — 28.5% |
| All | Hispanic/Latino<sup>3</sup> | M | 477.0 | 714.8 | 237.8 | 49.9% | 48.9% — 50.8% |
| All | Hispanic/Latino | F | 390.7 | 535.4 | 144.6 | 37.0% | 36.0% — 38.0% |
| All | White | M | 1,315.4 | 1514.6 | 199.2 | 15.1% | 14.9% — 15.4% |
| All | White | F | 1,219.3 | 1374.4 | 155.1 | 12.7% | 12.5% — 13.0% |
| 13-24 | All | All | 61.6 | 73.8 | 12.2 | 19.9% | 18.3% — 21.4% |
| 25-34 | All | All | 128.8 | 159.5 | 30.7 | 23.9% | 22.7% — 25.0% |
| 35-44 | All | All | 197.0 | 248.0 | 51.0 | 25.9% | 24.9% — 26.9% |
| 45-54 | All | All | 394.2 | 473.5 | 79.3 | 20.1% | 19.5% — 20.8% |
| 55-64 | All | All | 885.0 | 1038.9 | 154.0 | 17.4% | 17.0% — 17.8% |
| 65+ | All | All | 3,959.6 | 4508.5 | 548.9 | 13.9% | 13.7% — 14.0% |
| 65-74 | All | All | 1,773.8 | 2072.3 | 298.4 | 16.8% | 16.5% — 17.2% |
| 75-84 | All | All | 4,346.5 | 4997.0 | 650.5 | 15.0% | 14.7% — 15.3% |
| 85+ | All | All | 13,339.1 | 15210.9 | 1871.7 | 14.0% | 13.8% — 14.3% |

<sup>1</sup>Rates calculated by dividing deaths in a group by overall group population. All rates are per 100,000 population.
<sup>2</sup>Refers to sex determined at death. M denotes “Male,” F denotes “Female.”
<sup>3</sup> Hispanic/Latino persons can be of any race.

**Table 2:** Mortality rates^1^ among persons with diagnosed HIV (PWDH) aged ≥13 years, by age, race/ethnicity, and sex^2^, 2018–2020—United States

| Age | Race/Ethnicity | Sex<sup>2</sup> | Mortality Rate, 2018-19 | Mortality Rate, 2020 | Change from 2018-19 (Absolute) | Change from 2018-19 (%) | Change from 2018-19 (%), 95% confidence interval |
| --- | --- | --- | --- | --- | --- | --- | --- |
| All | All | All | 1,575.6 | 1722.7 | 147.1 | 9.3% | 7.4% – 11.3% |
| All | All | M | 1,560.3 | 1693.0 | 132.7 | 8.5% | 6.3% – 10.8% |
| All | All | F | 1,625.8 | 1821.5 | 195.7 | 12.0% | 8.0% – 16.2% |
| All | Black/African-American | All | 1,667.3 | 1847.3 | 180.0 | 10.8% | 7.8% – 13.9% |
| All | Hispanic/Latino | All | 1,178.7 | 1319.6 | 140.9 | 12.0% | 7.2% – 16.9% |
| All | White | All | 1,723.8 | 1802.6 | 78.8 | 4.6% | 1.2% – 8.0% |
| All | Black/African-American | M | 1,705.3 | 1891.0 | 185.7 | 10.9% | 7.3% – 14.6% |
| All | Black/African-American | F | 1,592.1 | 1759.2 | 167.0 | 10.5% | 5.3% – 16.0% |
| All | Hispanic/Latino<sup>3</sup> | M | 1,152.8 | 1268.5 | 115.7 | 10.0% | 4.8% – 15.5% |
| All | Hispanic/Latino | F | 1,290.6 | 1546.7 | 256.1 | 19.8% | 9.2% – 31.6% |
| All | White | M | 1,677.7 | 1767.3 | 89.6 | 5.3% | 1.7% – 9.1% |
| All | White | F | 2,030.1 | 2034.8 | 4.7 | 0.2% | -7.8% – 8.9% |
| 13-24 | All | All | 419.7 | 506.9 | 88.4 | 21.1% | -1.1% – 48.3% |
| 25-34 | All | All | 663.5 | 727.5 | 64.0 | 9.6% | 2.1% – 17.7% |
| 35-44 | All | All | 928.2 | 998.4 | 70.3 | 7.6% | 1.8% – 13.6% |
| 45-54 | All | All | 1,426.2 | 1429.2 | 3.0 | 0.2% | -3.6% – 4.2% |
| 55-64 | All | All | 2,089.8 | 2192.5 | 102.7 | 4.9% | 1.7% – 8.2% |
| 65+ | All | All | 3,531.0 | 3898.8 | 367.8 | 10.4% | 6.6% – 14.3% |

<sup>1</sup>Rates calculated by dividing deaths in a group by overall group population, in this case PWDH deaths divided by diagnosed HIV prevalence in the specified group. All rates are per 100,000 population.
<sup>2</sup>Refers to assigned sex at birth. M denotes “Male,” F denotes “Female.”
<sup>3</sup> Hispanic/Latino persons can be of any race.

Black and Hispanic/Latino populations showed larger increases in mortality rates from 2018-2019 to 2020, in both absolute and relative terms, compared to White populations, in the general population (295.2 and 29.7% for Black, 191.5 and 44.1% for Hispanic/Latino, 176.7 and 14.0% for White). Among PWDH, absolute increases were largest for Black and Hispanic/Latino PWH (180.0 for Black, 140.9 for Hispanic/Latino, 78.8 for White), while the relative changes were similar among the different racial/ethnic groups (confidence intervals overlap).

Absolute and relative mortality rate increases were larger among men in the general population (215.6 and 20.0% compared to 156.9 and 16.1% for women), however this was not clearly observed in the PWDH population (confidence intervals for the two rates overlap).

Among the entire populations, the absolute increase in mortality among PWDH was 21% smaller than the general population (an increase of 147.1 for PWDH compared to 185.6 for the general population). Absolute mortality increases were smaller for PWDH across each examined racial/ethnic group, and among all groups aged 45 and older. Among female PWDH (both overall and for each race/ethnicity), Hispanic/Latino PWDH, PWDH younger than 45, and PWDH aged 55-64, absolute mortality increases showed no statistically significant difference from corresponding increases in the general population.

We estimate approximately 1550 excess deaths occurred among PWDH in 2020 (Table 3). Excess deaths were particularly concentrated among Black PWDH and PWDH ≥55 years. All groups showed positive excess mortality besides White females, PWDH aged 13-24, and PWDH aged 45-54, for whom overall mortality levels were consistent with pre-pandemic trends. We note these numbers are consistent with those found in [18].

**Table 3:** Excess mortality^1^ among persons with diagnosed HIV (PWDH) aged ≥13 years, by age, race/ethnicity, and sex^2^, 2018–2020—United States

| Age | Race/ethnicity | Sex<sup>2</sup> | Excess Deaths, 2020 | 95% confidence interval |  |  |
| --- | --- | --- | --- | --- | --- | --- |
| All | All | All | 1,551 | 1,229 | — | 1,877 |
| All | All | M | 1,075 | 796 | — | 1,365 |
| All | All | F | 477 | 317 | — | 643 |
| All | Black/African-American | All | 772 | 558 | — | 995 |
| All | Hispanic/Latino<sup>3</sup> | All | 347 | 209 | — | 490 |
| All | White | All | 241 | 63 | — | 422 |
| All | Black/African-American | M | 533 | 357 | — | 714 |
| All | Black/African-American | F | 238 | 120 | — | 363 |
| All | Hispanic/Latino | M | 232 | 111 | — | 359 |
| All | Hispanic/Latino | F | 116 | 54 | — | 184 |
| All | White | M | 238 | 76 | — | 405 |
| All | White | F | 2 | -64 | — | 73 |
| 13-24 | All | All | 25 | -1 | — | 58 |
| 25-34 | All | All | 104 | 23 | — | 191 |
| 35-44 | All | All | 138 | 33 | — | 249 |
| 45-54 | All | All | 8 | -131 | — | 152 |
| 55-64 | All | All | 290 | 100 | — | 484 |
| 65+ | All | All | 475 | 301 | — | 652 |

<sup>1</sup>Excess mortality in a group is calculated by multiplying the total population size, the 2018-19 mortality rate, and the calculated percent change in the mortality rate in 2020 from 2018-19.
<sup>2</sup>Refers to assigned sex at birth. M denotes “Male,” F denotes “Female.”
<sup>3</sup> Hispanic/Latino persons can be of any race.

Statistically significant decreases in RR from 2018-2019 to 2020 were observed in most populations (Tables 4 and 5). RR drops were highest among Hispanic/Latino PWDH (particularly pronounced among males), Black males, and persons aged 45-54. Females, persons aged 13-24, and persons aged 65 and older showed no statistically significant change in RR (Table 5). No statistically significant increases in RR were found.

**Table 4:** Relative mortality risk^1^ among persons with diagnosed HIV (PWDH) aged ≥13 years, by age, race/ethnicity, and sex^2^, 2018–2020—United States

| Age | Race/ethnicity | Sex<sup>2</sup> | RR, 2018-19 | 95% Confidence interval |  |  | RR, 2020 | 95% Confidence interval |  |  |
| --- | --- | --- | --- | --- | --- | --- | --- | --- | --- | --- |
| All | All | All | 1.5 | 1.5 | — | 1.6 | 1.4 | 1.4 | — | 1.5 |
| All | All | M | 1.5 | 1.4 | — | 1.5 | 1.3 | 1.3 | — | 1.3 |
| All | All | F | 1.7 | 1.6 | — | 1.7 | 1.6 | 1.6 | — | 1.7 |
| All | Black/African-American | All | 1.7 | 1.7 | — | 1.7 | 1.4 | 1.4 | — | 1.5 |
| All | Hispanic/Latino<sup>3</sup> | All | 2.7 | 2.6 | — | 2.8 | 2.1 | 2.0 | — | 2.2 |
| All | White | All | 1.4 | 1.3 | — | 1.4 | 1.3 | 1.2 | — | 1.3 |
| All | Black/African-American | M | 1.6 | 1.5 | — | 1.6 | 1.3 | 1.3 | — | 1.4 |
| All | Black/African-American | F | 1.8 | 1.7 | — | 1.8 | 1.5 | 1.5 | — | 1.6 |
| All | Hispanic/Latino | M | 2.4 | 2.4 | — | 2.5 | 1.8 | 1.7 | — | 1.8 |
| All | Hispanic/Latino | F | 3.3 | 3.1 | — | 3.5 | 2.9 | 2.7 | — | 3.1 |
| All | White | M | 1.3 | 1.3 | — | 1.3 | 1.2 | 1.1 | — | 1.2 |
| All | White | F | 1.7 | 1.6 | — | 1.8 | 1.5 | 1.4 | — | 1.6 |
| 13-24 | All | All | 6.8 | 6.0 | — | 7.7 | 6.9 | 5.8 | — | 8.1 |
| 25-34 | All | All | 5.2 | 4.9 | — | 5.4 | 4.6 | 4.3 | — | 4.8 |
| 35-44 | All | All | 4.7 | 4.6 | — | 4.9 | 4.0 | 3.9 | — | 4.2 |
| 45-54 | All | All | 3.6 | 3.5 | — | 3.7 | 3.0 | 2.9 | — | 3.1 |
| 55-64 | All | All | 2.4 | 2.3 | — | 2.4 | 2.1 | 2.1 | — | 2.2 |
| 65+ | All | All | 0.9 | 0.9 | — | 0.9 | 0.9 | 0.8 | — | 0.9 |

<sup>1</sup>Relative mortality risk for a group is determined by dividing the group PWDH mortality rate by the rate among the general population group.
<sup>2</sup>Refers to assigned sex at birth among persons with diagnosed HIV and sex determined at death. M denotes “Male,” F denotes “Female.”
<sup>3</sup> Hispanic/Latino persons can be of any race.

**Table 5:** Change in relative mortality risk^1^ among persons with diagnosed HIV (PWDH) aged ≥13 years, by age, race/ethnicity, and sex^2^, 2018–2020—United States

| Age | Race/ethnicity | Sex<sup>2</sup> | RR2020/<br>RR2018-19 | 95% Confidence interval |  |  | p-value |
| --- | --- | --- | --- | --- | --- | --- | --- |
| All | All | All | 0.93 | 0.91 | — | 0.94 | 0.00 |
| All | All | M | 0.90 | 0.89 | — | 0.92 | 0.00 |
| All | All | F | 0.96 | 0.93 | — | 1.00 | 0.05 |
| All | Black/African-American | All | 0.85 | 0.83 | — | 0.88 | 0.00 |
| All | Hispanic/Latino<sup>3</sup> | All | 0.78 | 0.74 | — | 0.81 | 0.00 |
| All | White | All | 0.92 | 0.89 | — | 0.95 | 0.00 |
| All | Black/African-American | M | 0.84 | 0.82 | — | 0.87 | 0.00 |
| All | Black/African-American | F | 0.86 | 0.82 | — | 0.91 | 0.00 |
| All | Hispanic/Latino | M | 0.73 | 0.70 | — | 0.77 | 0.00 |
| All | Hispanic/Latino | F | 0.87 | 0.80 | — | 0.96 | 0.01 |
| All | White | M | 0.91 | 0.88 | — | 0.95 | 0.00 |
| All | White | F | 0.89 | 0.82 | — | 0.97 | 0.01 |
| 13-24 | All | All | 1.01 | 0.82 | — | 1.23 | 0.94 |
| 25-34 | All | All | 0.89 | 0.82 | — | 0.95 | 0.00 |
| 35-44 | All | All | 0.85 | 0.81 | — | 0.90 | 0.00 |
| 45-54 | All | All | 0.83 | 0.80 | — | 0.87 | 0.00 |
| 55-64 | All | All | 0.89 | 0.87 | — | 0.92 | 0.00 |
| 65+ | All | All | 0.97 | 0.94 | — | 1.00 | 0.09 |

<sup>1</sup>Change in relative mortality risk for a group is determined by dividing the relative mortality risk in 2020 by the relative mortality risk in 2018-19
<sup>2</sup>Refers to assigned sex at birth among persons with diagnosed HIV and sex determined at death. M denotes “Male,” F denotes “Female.”
<sup>3</sup> Hispanic/Latino persons can be of any race.

In appendices A, B, and C, we provide further analyses based on two-way stratifications considering age and sex (Appendix A) and age and race/ethnicity (Appendix B). Three-way stratification of age, sex, and race/ethnicity is given in Appendix C. The results do not generally differ from the trends shown here, with most groups showing decreases in RR. Only one statistically significant increase was observed, among females aged 65 and older.

## Discussion

The results suggest that PWDH were not disproportionately affected, at least in terms of mortality, by COVID-19 or its second-order effects in 2020. While mortality increased for PWDH in 2020 across nearly all age, sex, and racial/ethnic groups, such increases were usually lower than mortality increases in the general population. These findings are in line with several meta-analyses and clinical studies [7]–[10], suggesting that, after controlling for underlying differences in mortality between PWDH and the general population, HIV did not exert a significant influence on COVID-19 mortality. As this study considers all mortality causes, and not just direct COVID-19 mortality, we also find no direct evidence that second-order COVID-19 deaths were more frequent among the PWDH population, and in fact may have had a lesser effect.

The results of this analysis are useful for future analyses of HIV in the United States. For modeling, surveillance, and intervention efforts, having a clear understanding of how COVID-19 changed mortality patterns in PWDH, and how such changes may influence future PWDH mortality and transmission, is of critical importance. We believe the analysis given here provides compelling evidence that the increases in PWDH mortality are generally smaller than those in the overall population, and future changes in mortality among PWDH due to COVID-19 may be expected to follow this pattern.

The smaller relative increase in mortality rates among PWDH compared with the general population may be explained in part by the higher baseline mortality rates among PWDH, making the impact of COVID-19 proportionally less significant. While this is an important factor, it does not explain the entirety of the decreases in RR observed, as *absolute* mortality increases were generally smaller among PWDH than the general population, with the exceptions of females (driven primarily by Hispanic/Latino females), and persons less than 45 years old.

The reasons why such mortality increases were smaller among PWDH are not immediately obvious and there are likely many. However, the proportion of PWDH who received HIV medical care in 2020 remained similar to previous years [19]. In contrast, several studies suggest that access to health care during 2020 among general population was delayed or reduced [20], [21]. Our findings could also be explained in part by lower COVID-19 morbidity among PWDH; however, this is not supported by current literature [3], [22]. We do not believe vaccination played a significant role in the current analysis, as vaccines were not widely available during the considered time period.

Analysis of excess mortality among PWDH shows that nearly half of the excess mortality in 2020 occurred among PWDH over 55 years, and that over half of excess deaths occurred among the Black and Hispanic/Latino PWDH populations. Nonetheless, a substantial share of excess mortality among PWDH occurred among younger age groups, who may be particularly at risk for overdose deaths, which increased sharply in 2020 [2]. Going forward, it will be important to monitor mortality trends among these younger cohorts.

We acknowledge that the present study is limited in several ways. Examining overall mortality, as was done here, does not distinguish cause of death. While this has some advantages, as we are interested not only in direct COVID-19 deaths, but also second-order deaths due to the pandemic, it also presents some issues. Particularly, we acknowledge possible problems as explained. For instance, if direct COVID-19 mortality were disproportionately high among PWDH, but second-order mortality disproportionately low, these effects may mask each other. While this may not be a problem for backward-facing analyses of PWDH mortality (where overall mortality is more important), it may affect future projections, as the rates of direct and second-order deaths from COVID-19 may change. In such a case, the stable risk ratio shown here may no longer be reliable. While we acknowledge this possibility, we stress that we do not believe this is likely, given the conclusions of previous studies [7]–[10]. As such, we advocate for interpreting the present study as evidence that neither direct nor second-order COVID-19 mortality was elevated among PWDH in 2020. Our study period of 2018-2019 may also represent a limitation, as the small time window, in addition to considering the 2018-19 rate as an aggregate over two years, may not account for long-term trends. Also, an analysis with more detailed age structuring may help further understanding, particularly among the 65+ group.

Finally, we acknowledge that prevalence and mortality data for PWDH in Kansas and Mississippi (for 2019) and North Carolina, Kansas, South Carolina and Vermont (in 2020) are not complete.

Future studies should concentrate on exploring possible factors that may further explain our findings, and in particular: vaccine uptake and efficacy among PWDH, as well as prevalence of long-term COVID-19 related complications.

Although mortality rates increased among PWDH from 2018-2019 to 2020, these increases were generally smaller than those observed in the general population. This resulted in decreases in the relative mortality risk ratio. While the extent to which the RR changed varied across different populations, nearly all populations showed a decrease. Females and persons aged 13-24 and aged 65 and older showed no statistically significant change in RR. A statistically significant increase was only observed in one group (females aged 65 and older). Our findings indicate that PWDH do not show a particularly elevated mortality risk from COVID-19 or its indirect effects, when compared with the general population. These findings are significant, as COVID-19 will remain a public health concern for the foreseeable future, and future modeling and intervention planning must take this into account. While we emphasize that COVID-19 does nonetheless represent a serious health risk for PWDH, the mortality risk does not appear to differ from that of the overall population.

## Data Availability

All data produced in the present study are available upon reasonable request to the authors.

## Acknowledgements

All funding for this study was provided by the Centers for Disease Control and Prevention.

## Author contributions

A. Viguerie: conceptualization, statistical analysis, writing/revising, data collection/curation, organization. R. Song: statistical analysis. K. Bosh: writing/revising, organization, table design. C. Lyles: writing/revising, supervision. P. Farnham: writing/revising, supervision.

The authors declare no potential conflicts of interest.

## Appendix A: Two-way stratification, age and sex-at-birth

In this section, we examine the populations shown in the main body text, further stratified by age group and sex-at-birth. We report the mortality rates for the general population and PWDH population in Tables A1 and A2, respectively, total excess deaths in 2020 in Table A3, the mortality risk ratios in Table A4, and the change in mortality risk ratios in Table A5.

All groups in the general population showed increases in mortality. Relative mortality increases were generally larger among younger groups, however, older groups showed larger absolute mortality increases. Among PWDH, many of the examined age groups did not show statistically significant change in mortality rates during 2020. However, large relative and absolute increases in mortality were observed in male PWDH over 55 and female PWDH aged 13-34 and aged 65+. Older PWDH, both male and female, comprised the majority of excess mortality among PWDH during 2020.

Among both males and females aged 13-24, the relative risk of mortality among PWDH did not show a statistically significant change between 2019 and 2020. Additionally, females aged 25-34 and 55+ showed no statistically significant change in mortality risk between 2018-19 and 2020. We note that the relative mortality risk ratio among female PWDH increased slightly in 2020, the only statistically significant increase in mortality RR found in our analyses. Among the other examined groups, mortality risk showed statistically significant decreases among PWDH. These indicators provide further evidence for a lack of disproportionate mortality among PWDH, across age and sex-at-birth groups, during 2020.

**Table A1:**
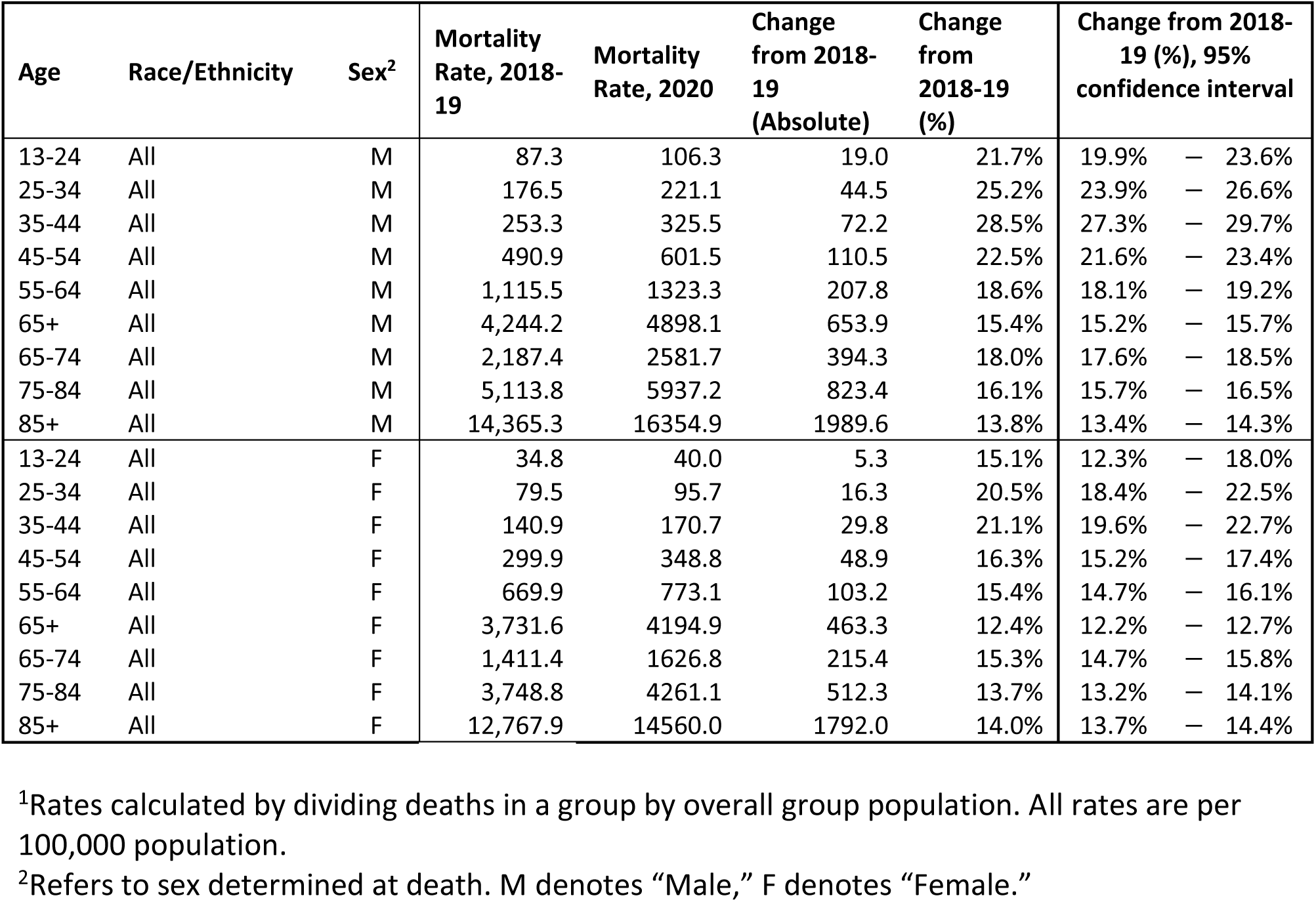
Mortality rates^1^ among the general population aged ≥13 years, stratified by age and sex^2^, 2018–2020—United States

**Table A2:** Mortality rates^1^ among persons with diagnosed HIV (PWDH) aged ≥13 years, stratified by age and sex^2^, 2018–2020—United States

| Age | Race/Ethnicity | Sex<sup>2</sup> | Mortality Rate, 2018-19 | Mortality Rate, 2020 | Change from 2018-19 (Absolute) | Change from 2018-19 (%) | Change from 2018-19 (%), 95% confidence interval |  |  |
| --- | --- | --- | --- | --- | --- | --- | --- | --- | --- |
| 13-24 | All | M | 408.2 | 486.4 | 78.2 | 19.2% | -5.4% | — | 50.1% |
| 25-34 | All | M | 633.7 | 668.7 | 35.0 | 5.5% | -2.6% | — | 14.4% |
| 35-44 | All | M | 873.9 | 954.4 | 80.5 | 9.2% | 2.3% | — | 16.5% |
| 45-54 | All | M | 1,389.8 | 1405.1 | 15.3 | 1.1% | -3.4% | — | 5.9% |
| 55-64 | All | M | 2,094.1 | 2197.7 | 103.6 | 4.9% | 1.3% | — | 8.7% |
| 65+ | All | M | 3,661.7 | 3927.6 | 265.9 | 7.3% | 3.1% | — | 11.6% |
| 13-24 | All | F | 457.7 | 588.1 | 130.4 | 28.5% | -15.5% | — | 95.5% |
| 25-34 | All | F | 809.2 | 1059.3 | 250.1 | 30.9% | 12.7% | — | 52.0% |
| 35-44 | All | F | 1,079.5 | 1132.7 | 53.3 | 4.9% | -5.2% | — | 16.2% |
| 45-54 | All | F | 1,531.8 | 1495.5 | -36.3 | -2.4% | -9.3% | — | 5.1% |
| 55-64 | All | F | 2,075.6 | 2175.2 | 99.6 | 4.8% | -1.7% | — | 11.7% |
| 65+ | All | F | 3,092.9 | 3803.4 | 710.5 | 23.0% | 14.1% | — | 32.6% |

<sup>1</sup>Rates calculated by dividing deaths in a group by overall group population, in this case PWDH deaths divided by diagnosed HIV prevalence in the specified group. All rates are per 100,000 population.
<sup>2</sup>Refers to assigned sex at birth. M denotes “Male,” F denotes “Female.”

**Table A3:** Excess mortality^1^ among persons with diagnosed HIV (PWDH) aged ≥13 years, stratified by age and sex^2^, 2018–2020—United States

| Age | Race/ethnicity | Sex<sup>2</sup> | Excess Deaths, 2020 | 95% confidence interval |  |  |
| --- | --- | --- | --- | --- | --- | --- |
| 13-24 | All | M | 18 | -5 | — | 47 |
| 25-34 | All | M | 47 | -22 | — | 124 |
| 35-44 | All | M | 119 | 30 | — | 214 |
| 45-54 | All | M | 29 | -88 | — | 153 |
| 55-64 | All | M | 225 | 59 | — | 395 |
| 65+ | All | M | 264 | 113 | — | 421 |
| 13-24 | All | F | 8 | -4 | — | 25 |
| 25-34 | All | F | 65 | 27 | — | 110 |
| 35-44 | All | F | 26 | -26 | — | 85 |
| 45-54 | All | F | -25 | -97 | — | 53 |
| 55-64 | All | F | 65 | -23 | — | 160 |
| 65+ | All | F | 213 | 131 | — | 302 |

<sup>1</sup>Excess mortality in a group is calculated by multiplying the total population size, the 2018-19 mortality rate, and the calculated percent change in the mortality rate in 2020 from 2018-19.
<sup>2</sup>Refers to assigned sex at birth. M denotes “Male,” F denotes “Female.”

**Table A4:**
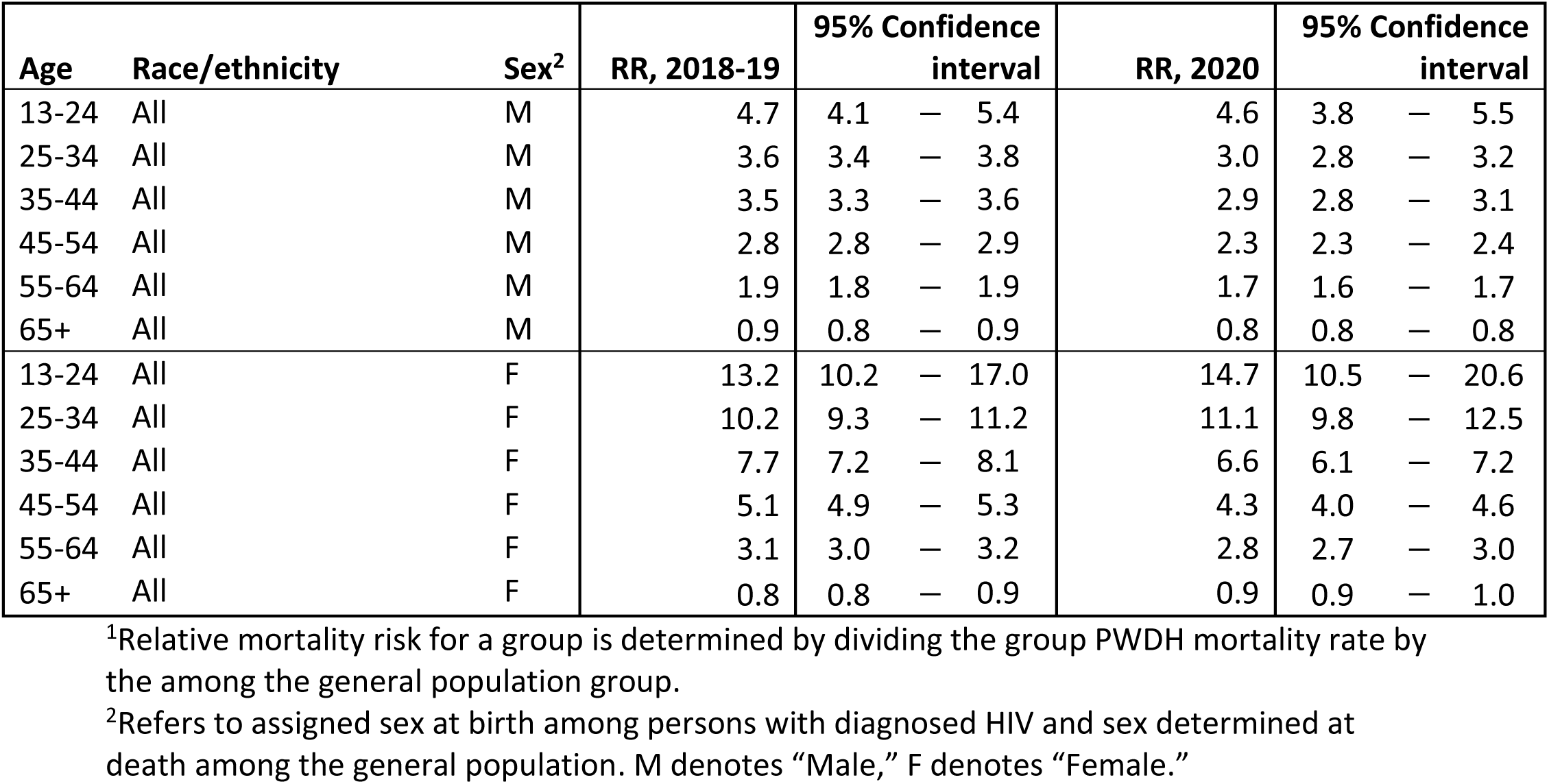
Relative mortality risk^1^ among persons with diagnosed HIV (PWDH) aged ≥13 years, stratified by age and sex^2^, 2018–2020—United States

**Table A5:** Change relative mortality risk^1^ among persons with diagnosed HIV (PWDH) aged ≥13 years, stratified by age sex^2^, 2018–2020—United States

| Age | Race/ethnicity | Sex<sup>2</sup> | RR2018-19/RR2020 | 95% Confidence interval |  |  | p-value |
| --- | --- | --- | --- | --- | --- | --- | --- |
| 13-24 | All | M | 0.98 | 0.78 | — | 1.23 | 0.86 |
| 25-34 | All | M | 0.84 | 0.78 | — | 0.91 | 0.00 |
| 35-44 | All | M | 0.85 | 0.80 | — | 0.91 | 0.00 |
| 45-54 | All | M | 0.83 | 0.79 | — | 0.86 | 0.00 |
| 55-64 | All | M | 0.88 | 0.85 | — | 0.92 | 0.00 |
| 65+ | All | M | 0.93 | 0.89 | — | 0.97 | 0.00 |
| 13-24 | All | F | 1.12 | 0.73 | — | 1.70 | 0.61 |
| 25-34 | All | F | 1.09 | 0.93 | — | 1.26 | 0.28 |
| 35-44 | All | F | 0.87 | 0.78 | — | 0.96 | 0.01 |
| 45-54 | All | F | 0.84 | 0.78 | — | 0.90 | 0.00 |
| 55-64 | All | F | 0.91 | 0.85 | — | 0.97 | 0.00 |
| 65+ | All | F | 1.09 | 1.01 | — | 1.18 | 0.02 |

<sup>1</sup>Change in relative mortality risk for a group is determined by dividing the relative mortality risk in 2020 by the relative mortality risk in 2018-19
<sup>2</sup>Refers to assigned sex at birth among persons with diagnosed HIV and sex determined at death. M denotes “Male,” F denotes “Female.”

## Appendix B: Two-way stratification, age and race/ethnicity

In this appendix, we examine the populations shown in the main body text, further stratified by age group and race/ethnicity (Black, Hispanic/Latino, and White). We report the mortality rates for the general population and PWDH population in Tables B1 and B2, respectively, total excess deaths among PWDH in 2020 in Table B3, the mortality risk ratios in Table B4, and the change in mortality risk ratios in Table B5.

All age groups in all racial/ethnic groups showed increased mortality among the general population, with both absolute and relative increases significantly higher for Black and Hispanic/Latino populations, as compared to White populations, across all age groups. Among PWDH, statistically significant mortality increases generally occurred among the individuals in the Black and Hispanic/Latino population. Increases in mortality were less pronounced among White PWDH, with White PWDH aged 65 and older showing no significant change in mortality in 2020.

The relative risk of mortality among PWDH aged 13-24 did not show a statistically significant change between 2018-19 and 2020 across each examined race/ethnicity. Among the other examined groups, mortality risk showed statistically significant decreases among PWDH. These indicators provide further evidence for a lack of disproportionate mortality among PWDH, across racial/ethnic and age groups, during 2020. In fact, the data suggests that PWDH showed a smaller increase in mortality susceptibility as compared to the general population.

**Table B1:**
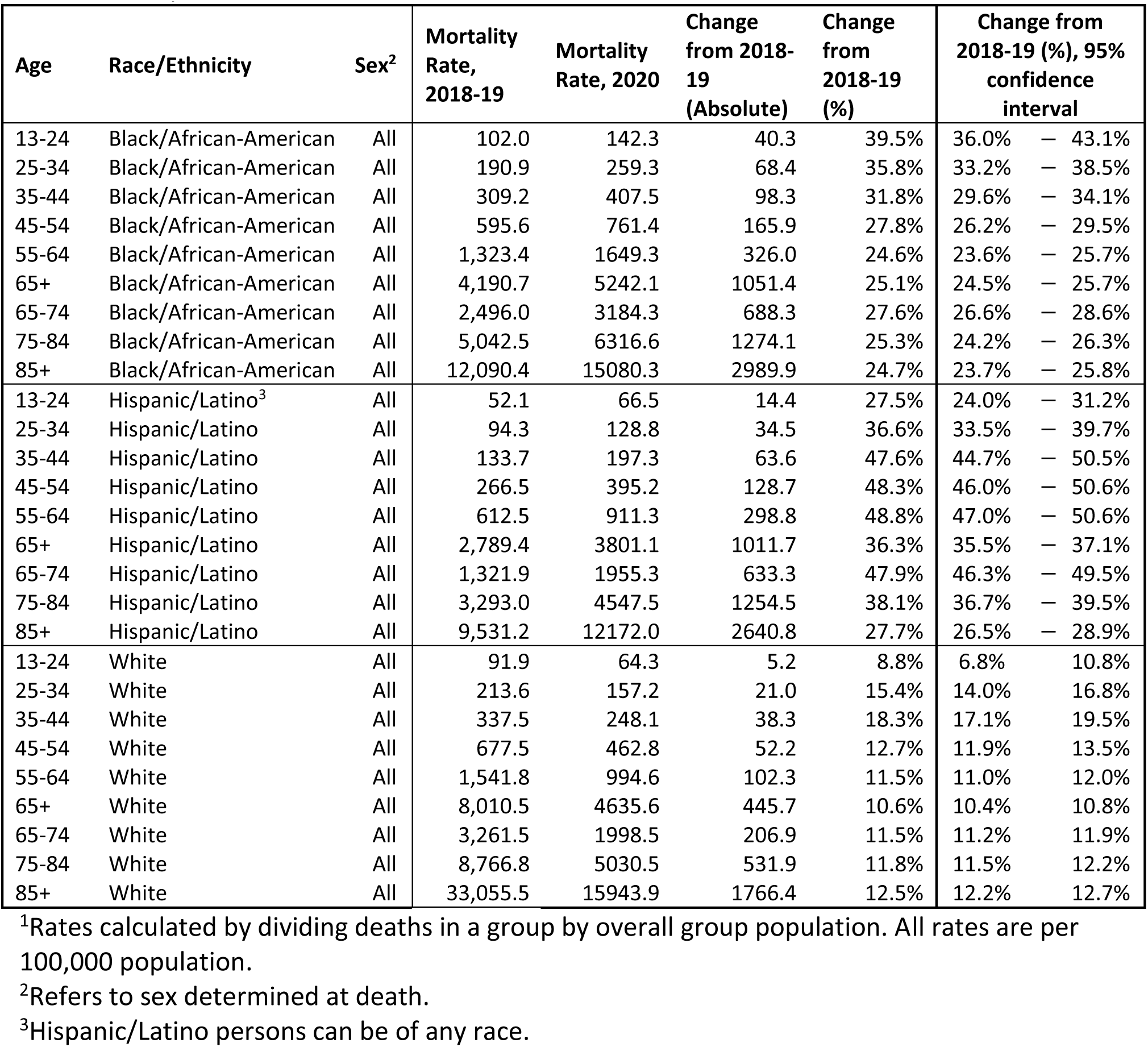
Mortality rates^1^ among the general population aged ≥13 years, stratified by age and race/ethnicity, 2018–2020—United States

**Table B2:** Mortality rates^1^ among persons with diagnosed HIV (PWDH) aged ≥13 years, stratified by age and race/ethnicity, 2018–2020—United States

| Age | Race/Ethnicity | Sex<sup>2</sup> | Mortality Rate, 2018-19 | Mortality Rate, 2020 | Change from 2018-19 (Absolute) | Change from 2018-19 (%) | Change from 2018-19 (%), 95% confidence interval |
| --- | --- | --- | --- | --- | --- | --- | --- |
| 13-24 | Black/African-American | All | 423.2 | 512.5 | 89.3 | 21.1% | -7.3% — 58.3% |
| 25-34 | Black/African-American | All | 713.2 | 728.2 | 15.0 | 2.1% | -7.6% — 12.9% |
| 35-44 | Black/African-American | All | 1,012.2 | 1132.7 | 120.5 | 11.9% | 3.2% — 21.3% |
| 45-54 | Black/African-American | All | 1,571.6 | 1569.1 | -2.5 | -0.2% | -6.0% — 6.0% |
| 55-64 | Black/African-American | All | 2,352.3 | 2506.7 | 154.4 | 6.6% | 1.7% — 11.7% |
| 65+ | Black/African-American | All | 3,837.2 | 4402.5 | 565.2 | 14.7% | 8.8% — 21.0% |
| 13-24 | Hispanic/Latino<sup>3</sup> | All | 270.9 | 376.7 | 105.8 | 39.1% | -15.5% — 128.8% |
| 25-34 | Hispanic/Latino | All | 449.5 | 474.2 | 24.7 | 5.5% | -11.3% — 25.5% |
| 35-44 | Hispanic/Latino | All | 565.5 | 581.6 | 16.2 | 2.9% | -10.2% — 17.8% |
| 45-54 | Hispanic/Latino | All | 828.0 | 875.8 | 47.8 | 5.8% | -4.4% — 17.0% |
| 55-64 | Hispanic/Latino | All | 1,371.2 | 1433.4 | 62.2 | 4.5% | -4.1% — 14.0% |
| 65+ | Hispanic/Latino | All | 2,452.1 | 2787.5 | 335.4 | 13.7% | 3.0% — 25.5% |
| 13-24 | White | All | 578.6 | 574.1 | -4.5 | -0.8% | -39.9% — 63.8% |
| 25-34 | White | All | 743.7 | 910.2 | 166.5 | 22.4% | 5.3% — 42.3% |
| 35-44 | White | All | 1,007.7 | 1087.3 | 79.6 | 7.9% | -3.2% — 20.3% |
| 45-54 | White | All | 1,494.6 | 1457.6 | -37.0 | -2.5% | -9.1% — 4.7% |
| 55-64 | White | All | 1,922.1 | 1921.2 | -1.0 | -0.1% | -5.3% — 5.5% |
| 65+ | White | All | 3,313.5 | 3367.9 | 54.4 | 1.6% | -4.3% — 7.9% |

<sup>1</sup>Rates calculated by dividing deaths in a group by overall group population, in this case PWDH deaths divided by diagnosed HIV prevalence in the specified group. All rates are per 100,000 population.
<sup>2</sup>Refers to sex determined assigned at birth.
<sup>3</sup> Hispanic/Latino persons can be of any race.

**Table B3:**
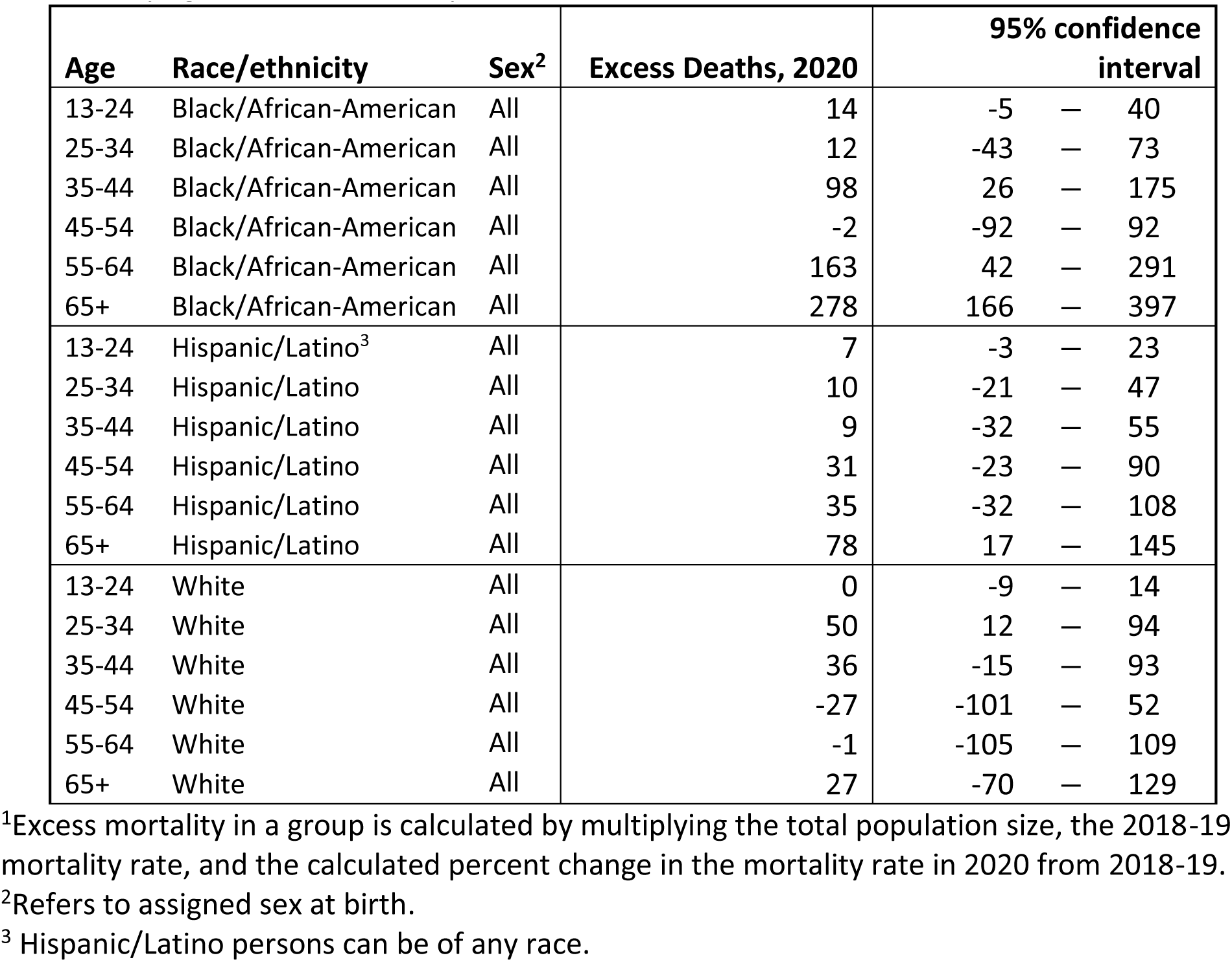
Excess mortality^1^ among persons with diagnosed HIV (PWDH) aged ≥13 years, stratified by age and race/ethnicity, 2018–2020—United States

| Age | Race/ethnicity | Sex<sup>2</sup> | Excess Deaths, 2020 | 95% confidence interval |  |  |
| --- | --- | --- | --- | --- | --- | --- |
| 13-24 | Black/African-American | All | 14 | -5 | — | 40 |
| 25-34 | Black/African-American | All | 12 | -43 | — | 73 |
| 35-44 | Black/African-American | All | 98 | 26 | — | 175 |
| 45-54 | Black/African-American | All | -2 | -92 | — | 92 |
| 55-64 | Black/African-American | All | 163 | 42 | — | 291 |
| 65+ | Black/African-American | All | 278 | 166 | — | 397 |
| 13-24 | Hispanic/Latino<sup>3</sup> | All | 7 | -3 | — | 23 |
| 25-34 | Hispanic/Latino | All | 10 | -21 | — | 47 |
| 35-44 | Hispanic/Latino | All | 9 | -32 | — | 55 |
| 45-54 | Hispanic/Latino | All | 31 | -23 | — | 90 |
| 55-64 | Hispanic/Latino | All | 35 | -32 | — | 108 |
| 65+ | Hispanic/Latino | All | 78 | 17 | — | 145 |
| 13-24 | White | All | 0 | -9 | — | 14 |
| 25-34 | White | All | 50 | 12 | — | 94 |
| 35-44 | White | All | 36 | -15 | — | 93 |
| 45-54 | White | All | -27 | -101 | — | 52 |
| 55-64 | White | All | -1 | -105 | — | 109 |
| 65+ | White | All | 27 | -70 | — | 129 |

<sup>1</sup>Excess mortality in a group is calculated by multiplying the total population size, the 2018-19 mortality rate, and the calculated percent change in the mortality rate in 2020 from 2018-19.
<sup>2</sup>Refers to assigned sex at birth.
<sup>3</sup> Hispanic/Latino persons can be of any race.

**Table B4:** Relative mortality risk^1^ among persons with diagnosed HIV (PWDH) aged ≥13 years, stratified by age and race/ethnicity, 2018–2020—United States

| Age | Race/ethnicity | Sex<sup>2</sup> | RR, 2018-19 | 95% Confidence interval |  |  | RR, 2020 | 95% Confidence interval |  |  |
| --- | --- | --- | --- | --- | --- | --- | --- | --- | --- | --- |
| 13-24 | Black/African-American | All | 4.2 | 3.5 | — | 4.9 | 3.6 | 2.9 | — | 4.5 |
| 25-34 | Black/African-American | All | 3.7 | 3.5 | — | 4.0 | 2.8 | 2.6 | — | 3.1 |
| 35-44 | Black/African-American | All | 3.3 | 3.1 | — | 3.4 | 2.8 | 2.6 | — | 3.0 |
| 45-54 | Black/African-American | All | 2.6 | 2.6 | — | 2.7 | 2.1 | 2.0 | — | 2.2 |
| 55-64 | Black/African-American | All | 1.8 | 1.7 | — | 1.8 | 1.5 | 1.5 | — | 1.6 |
| 65+ | Black/African-American | All | 0.9 | 0.9 | — | 1.0 | 0.8 | 0.8 | — | 0.9 |
| 13-24 | Hispanic/Latino<sup>3</sup> | All | 5.2 | 3.8 | — | 7.1 | 5.7 | 3.8 | — | 8.4 |
| 25-34 | Hispanic/Latino | All | 4.8 | 4.3 | — | 5.3 | 3.7 | 3.2 | — | 4.2 |
| 35-44 | Hispanic/Latino | All | 4.2 | 3.9 | — | 4.6 | 3.0 | 2.6 | — | 3.3 |
| 45-54 | Hispanic/Latino | All | 3.1 | 2.9 | — | 3.3 | 2.2 | 2.0 | — | 2.4 |
| 55-64 | Hispanic/Latino | All | 2.2 | 2.1 | — | 2.4 | 1.6 | 1.5 | — | 1.7 |
| 65+ | Hispanic/Latino | All | 0.9 | 0.8 | — | 0.9 | 0.7 | 0.7 | — | 0.8 |
| 13-24 | White | All | 9.8 | 7.4 | — | 12.9 | 8.9 | 5.9 | — | 13.5 |
| 25-34 | White | All | 5.5 | 5.0 | — | 6.0 | 5.8 | 5.1 | — | 6.5 |
| 35-44 | White | All | 4.8 | 4.5 | — | 5.1 | 4.4 | 4.0 | — | 4.8 |
| 45-54 | White | All | 3.6 | 3.5 | — | 3.8 | 3.2 | 3.0 | — | 3.3 |
| 55-64 | White | All | 2.2 | 2.1 | — | 2.2 | 1.9 | 1.9 | — | 2.0 |
| 65+ | White | All | 0.8 | 0.8 | — | 0.8 | 0.7 | 0.7 | — | 0.8 |

<sup>1</sup>Relative mortality risk for a group is determined by dividing the group PWDH mortality rate by the among the general population group.
<sup>2</sup>Refers to assigned sex at birth among persons with diagnosed HIV and sex determined at death among the general population.
<sup>3</sup> Hispanic/Latino persons can be of any race.

**Table B5:** Change relative mortality risk^1^ among persons with diagnosed HIV (PWDH) aged ≥13 years, stratified by age race/ethnicity, 2018–2020—United States

| Age | Race/ethnicity | Sex<sup>2</sup> | RR2018-19/RR2020 | 95% Confidence interval |  |  | p-value |
| --- | --- | --- | --- | --- | --- | --- | --- |
| 13-24 | Black/African-American | All | 0.87 | 0.66 | — | 1.14 | 0.30 |
| 25-34 | Black/African-American | All | 0.75 | 0.68 | — | 0.83 | 0.00 |
| 35-44 | Black/African-American | All | 0.85 | 0.78 | — | 0.92 | 0.00 |
| 45-54 | Black/African-American | All | 0.78 | 0.73 | — | 0.83 | 0.00 |
| 55-64 | Black/African-American | All | 0.86 | 0.82 | — | 0.90 | 0.00 |
| 65+ | Black/African-American | All | 0.92 | 0.87 | — | 0.97 | 0.00 |
| 13-24 | Hispanic/Latino<sup>3</sup> | All | 1.09 | 0.66 | — | 1.80 | 0.73 |
| 25-34 | Hispanic/Latino | All | 0.77 | 0.65 | — | 0.92 | 0.00 |
| 35-44 | Hispanic/Latino | All | 0.70 | 0.61 | — | 0.80 | 0.00 |
| 45-54 | Hispanic/Latino | All | 0.71 | 0.64 | — | 0.79 | 0.00 |
| 55-64 | Hispanic/Latino | All | 0.70 | 0.64 | — | 0.77 | 0.00 |
| 65+ | Hispanic/Latino | All | 0.83 | 0.76 | — | 0.92 | 0.00 |
| 13-24 | White | All | 0.91 | 0.55 | — | 1.51 | 0.72 |
| 25-34 | White | All | 1.07 | 0.92 | — | 1.24 | 0.40 |
| 35-44 | White | All | 0.91 | 0.82 | — | 1.02 | 0.10 |
| 45-54 | White | All | 0.87 | 0.81 | — | 0.93 | 0.00 |
| 55-64 | White | All | 0.90 | 0.85 | — | 0.95 | 0.00 |
| 65+ | White | All | 0.92 | 0.87 | — | 0.98 | 0.01 |

<sup>1</sup>Change in relative mortality risk for a group is determined by dividing the relative mortality risk in 2020 by the relative mortality risk in 2018-19
<sup>2</sup>Refers to assigned sex at birth among persons with diagnosed HIV and sex determined at death. M denotes “Male,” F denotes “Female.”
<sup>3</sup> Hispanic/Latino persons can be of any race.

## Appendix C: Age, sex, race/ethnicity 3-way stratification

In this appendix, we examine the populations shown in the main body text, further stratified by age group, sex at birth, and race/ethnicity (Black, Hispanic/Latino, and White). We report the mortality rates for the general Black population and Black PWDH population in Tables C1 and C2, excess PWDH deaths in 2020 in Table C3, with the corresponding risk ratios in Table C4 and the changes in these ratios in Table C5. Data for the Hispanic/Latino and White populations are similarly reported in Tables C6-C10 and C11-C15, respectively.

Among the general population, both absolute and relative mortality increases were larger among Black and Hispanic/Latino populations as compared to White populations. In all racial/ethnic groups, men showed higher relative and absolute mortality increases as compared to women. Black and Hispanic/Latino PWDH showed particularly high relative and absolute mortality increases. Relative and absolute mortality increases were large among young (under 34) and older (65+) female PWDH of all racial/ethnic groups.

The relative risk of mortality among PWDH aged 13-24 did not show a statistically significant change between 2018-19 and 2020 across each examined race/ethnicity and sex-at-birth group. Additionally, females aged 25-34 showed no statistically significant change in mortality risk across all racial/ethnic groups. Hispanic/Latino female PWDH aged 13-34 and 45+ showed no changes in mortality risk from 2018-19 to 2020. Finally, White PWDH, except for males aged 45+ and females aged 45-54, did not show changes mortality risk from 2018-19 - 2020.

Among the other examined groups, mortality risk showed statistically significant decreases among PWDH. These indicators provide further evidence for a lack of disproportionate mortality among PWDH, across racial/ethnic and age groups, during 2020. These decreases tended to be more common among male, older PWDH, and Black/Latino PWDH. Persons in these groups showed increased mortality susceptibility in the among the general population, increases which were smaller among the corresponding PWDH population. These data further support the conclusion that PWDH showed either no change, or slight decreases in, mortality risk during 2020. Further, this remained true across different age, racial/ethnic, and sex-at-birth, groups.

We remark that, in this analysis, some groups showed fewer than 12 deaths in certain years. Such groups are indicated in red in the following tables.

**Table C1:** Mortality rates^1^ among the general Black/African-American population aged ≥13 years, stratified by age and sex^2^, 2018–2020—United States

| Age | Race/Ethnicity | Sex<sup>2</sup> | Mortality Rate, 2018-19 | Mortality Rate, 2020 | Change from 2018-19 (Absolute) | Change from 2018-19 (%) | Change from 2018-19 (%), 95% confidence interval |
| --- | --- | --- | --- | --- | --- | --- | --- |
| 13-24 | Black/African-American | M | 153.9 | 215.4 | 61.5 | 39.9% | 35.9% – 44.1% |
| 25-34 | Black/African-American | M | 270.6 | 370.3 | 99.7 | 36.8% | 33.7% – 40.0% |
| 35-44 | Black/African-American | M | 405.3 | 539.7 | 134.4 | 33.2% | 30.3% – 36.1% |
| 45-54 | Black/African-American | M | 748.2 | 980.3 | 232.1 | 31.0% | 28.9% – 33.2% |
| 55-64 | Black/African-American | M | 1,691.9 | 2132.8 | 440.9 | 26.1% | 24.7% – 27.5% |
| 65+ | Black/African-American | M | 4,800.6 | 6084.1 | 1283.6 | 26.7% | 25.9% – 27.6% |
| 65-74 | Black/African-American | M | 3,238.5 | 4171.7 | 933.3 | 28.8% | 27.5% – 30.2% |
| 75-84 | Black/African-American | M | 6,234.7 | 7864.4 | 1629.7 | 26.1% | 24.7% – 27.6% |
| 85+ | Black/African-American | M | 13,200.8 | 16563.2 | 3362.4 | 25.5% | 23.7% – 27.3% |
| 13-24 | Black/African-American | F | 48.7 | 67.4 | 18.7 | 38.4% | 31.4% – 45.9% |
| 25-34 | Black/African-American | F | 112.1 | 148.5 | 36.3 | 32.4% | 27.7% – 37.3% |
| 35-44 | Black/African-American | F | 222.4 | 287.3 | 64.9 | 29.2% | 25.6% – 32.9% |
| 45-54 | Black/African-American | F | 461.1 | 569.2 | 108.0 | 23.4% | 21.0% – 25.9% |
| 55-64 | Black/African-American | F | 1,012.1 | 1240.4 | 228.2 | 22.6% | 20.9% – 24.2% |
| 65+ | Black/African-American | F | 3,776.7 | 4668.6 | 891.8 | 23.6% | 22.8% – 24.4% |
| 65-74 | Black/African-American | F | 1,938.1 | 2443.3 | 505.2 | 26.1% | 24.6% – 27.6% |
| 75-84 | Black/African-American | F | 4,291.4 | 5336.2 | 1044.8 | 24.3% | 22.9% – 25.8% |
| 85+ | Black/African-American | F | 11,606.8 | 14424.7 | 2817.8 | 24.3% | 23.0% – 25.6% |

<sup>1</sup>Rates calculated by dividing deaths in a group by overall group population. All rates are per 100,000 population.
<sup>2</sup>Refers to sex determined at death. M denotes “Male,” F denotes “Female.”

**Table C2:** Mortality rates^1^ among Black/African-American persons with diagnosed HIV (PWDH) aged ≥13 years, stratified by age and sex^2^, 2018–2020—United States

| Age | Race/Ethnicity | Sex<sup>2</sup> | Mortality Rate, 2018-19 | Mortality Rate, 2020 | Change from 2018-19 (Absolute) | Change from 2018-19 (%) | Change from 2018-19 (%), 95% confidence interval |  |  |
| --- | --- | --- | --- | --- | --- | --- | --- | --- | --- |
| 13-24 | Black/African-American | M | 420.4 | 505.1 | 84.7 | 20.2% | -11.5% | — | 63.1% |
| 25-34 | Black/African-American | M | 708.2 | 683.3 | -24.9 | -3.5% | -14.0% | — | 8.2% |
| 35-44 | Black/African-American | M | 991.6 | 1145.0 | 153.4 | 15.5% | 4.4% | — | 27.8% |
| 45-54 | Black/African-American | M | 1,630.0 | 1664.4 | 34.3 | 2.1% | -5.4% | — | 10.2% |
| 55-64 | Black/African-American | M | 2,500.4 | 2727.0 | 226.7 | 9.1% | 3.1% | — | 15.4% |
| 65+ | Black/African-American | M | 4,178.0 | 4694.5 | 516.5 | 12.4% | 5.4% | — | 19.8% |
| 13-24 | Black/African-American | F | 432.5 | 539.2 | 106.6 | 24.7% | -28.6% | — | 117.6% |
| 25-34 | Black/African-American | F | 732.6 | 913.8 | 181.2 | 24.7% | 1.3% | — | 53.5% |
| 35-44 | Black/African-American | F | 1,045.6 | 1110.2 | 64.6 | 6.2% | -7.1% | — | 21.4% |
| 45-54 | Black/African-American | F | 1,482.2 | 1431.3 | -50.9 | -3.4% | -12.5% | — | 6.6% |
| 55-64 | Black/African-American | F | 2,078.9 | 2109.7 | 30.8 | 1.5% | -6.8% | — | 10.5% |
| 65+ | Black/African-American | F | 3,184.7 | 3853.6 | 668.9 | 21.0% | 9.6% | — | 33.6% |

<sup>1</sup>Rates calculated by dividing deaths in a group by overall group population, in this case PWDH deaths divided by diagnosed HIV prevalence in the specified group. All rates are per 100,000 population.
<sup>2</sup>Refers to assigned sex at birth. M denotes “Male,” F denotes “Female.”

**Table C3:** Excess mortality^1^ among Black/African-American persons with diagnosed HIV (PWDH) aged ≥13 years, stratified by age and sex^2^, 2018–2020—United States

| Age | Race/ethnicity | Sex<sup>2</sup> | Excess Deaths, 2020 | 95% confidence interval |  |  |
| --- | --- | --- | --- | --- | --- | --- |
| 13-24 | Black/African-American | M | 11 | -6 | — | 34 |
| 25-34 | Black/African-American | M | -16 | -63 | — | 37 |
| 35-44 | Black/African-American | M | 81 | 23 | — | 145 |
| 45-54 | Black/African-American | M | 20 | -51 | — | 96 |
| 55-64 | Black/African-American | M | 154 | 53 | — | 262 |
| 65+ | Black/African-American | M | 365 | 225 | — | 510 |
| 13-24 | Black/African-American | F | 4 | -4 | — | 18 |
| 25-34 | Black/African-American | F | 28 | 1 | — | 60 |
| 35-44 | Black/African-American | F | 19 | -21 | — | 64 |
| 45-54 | Black/African-American | F | -20 | -74 | — | 39 |
| 55-64 | Black/African-American | F | 12 | -53 | — | 82 |
| 65+ | Black/African-American | F | 143 | 52 | — | 240 |

<sup>1</sup>Excess mortality in a group is calculated by multiplying the total population size, the 2018-19 mortality rate, and the calculated percent change in the mortality rate in 2020 from 2018-19.
<sup>2</sup>Refers to assigned sex at birth. M denotes “Male,” F denotes “Female.”

**Table C4:**
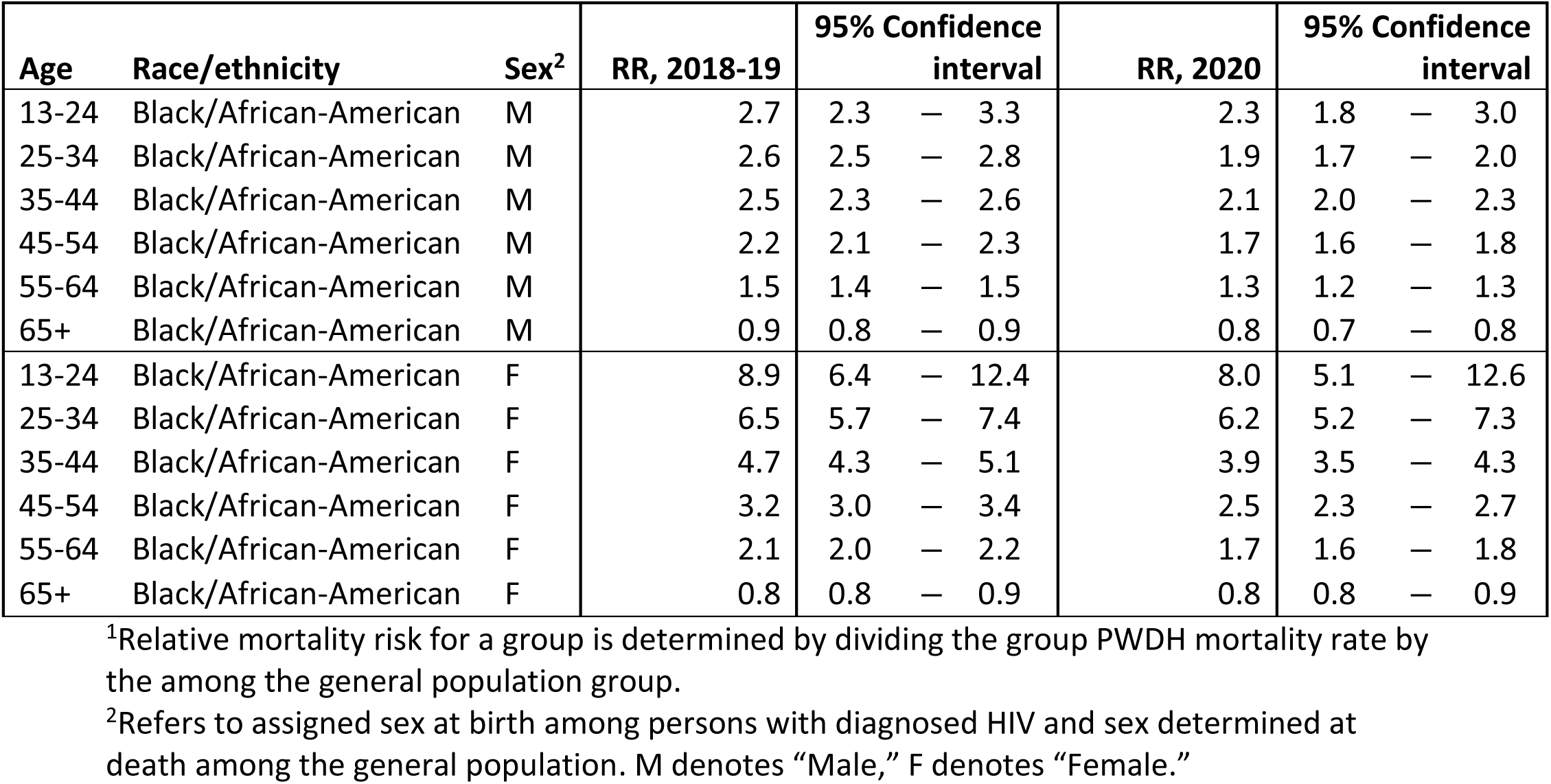
Relative mortality risk^1^ among Black/African-American persons with diagnosed HIV (PWDH) aged ≥13 years, stratified by age and sex^2^, 2018–2020—United States

| Age | Race/ethnicity | Sex<sup>2</sup> | RR, 2018-19 | 95% Confidence interval |  |  | RR, 2020 | 95% Confidence interval |  |  |
| --- | --- | --- | --- | --- | --- | --- | --- | --- | --- | --- |
| 13-24 | Black/African-American | M | 2.7 | 2.3 | — | 3.3 | 2.3 | 1.8 | — | 3.0 |
| 25-34 | Black/African-American | M | 2.6 | 2.5 | — | 2.8 | 1.9 | 1.7 | — | 2.0 |
| 35-44 | Black/African-American | M | 2.5 | 2.3 | — | 2.6 | 2.1 | 2.0 | — | 2.3 |
| 45-54 | Black/African-American | M | 2.2 | 2.1 | — | 2.3 | 1.7 | 1.6 | — | 1.8 |
| 55-64 | Black/African-American | M | 1.5 | 1.4 | — | 1.5 | 1.3 | 1.2 | — | 1.3 |
| 65+ | Black/African-American | M | 0.9 | 0.8 | — | 0.9 | 0.8 | 0.7 | — | 0.8 |
| 13-24 | Black/African-American | F | 8.9 | 6.4 | — | 12.4 | 8.0 | 5.1 | — | 12.6 |
| 25-34 | Black/African-American | F | 6.5 | 5.7 | — | 7.4 | 6.2 | 5.2 | — | 7.3 |
| 35-44 | Black/African-American | F | 4.7 | 4.3 | — | 5.1 | 3.9 | 3.5 | — | 4.3 |
| 45-54 | Black/African-American | F | 3.2 | 3.0 | — | 3.4 | 2.5 | 2.3 | — | 2.7 |
| 55-64 | Black/African-American | F | 2.1 | 2.0 | — | 2.2 | 1.7 | 1.6 | — | 1.8 |
| 65+ | Black/African-American | F | 0.8 | 0.8 | — | 0.9 | 0.8 | 0.8 | — | 0.9 |

<sup>1</sup>Relative mortality risk for a group is determined by dividing the group PWDH mortality rate by the among the general population group.
<sup>2</sup>Refers to assigned sex at birth among persons with diagnosed HIV and sex determined at death among the general population. M denotes “Male,” F denotes “Female.”

**Table C5:** Change relative mortality risk^1^ among Black/African-American persons with diagnosed HIV (PWDH) aged ≥13 years, stratified by age and sex^2^, 2018–2020—United States

| Age | Race/ethnicity | Sex<sup>2</sup> | RR2018-19/RR2020 | 95% Confidence interval |  |  | p-value |
| --- | --- | --- | --- | --- | --- | --- | --- |
| 13-24 | Black/African-American | M | 0.86 | 0.63 | — | 1.17 | 0.33 |
| 25-34 | Black/African-American | M | 0.71 | 0.63 | — | 0.79 | 0.00 |
| 35-44 | Black/African-American | M | 0.87 | 0.78 | — | 0.96 | 0.01 |
| 45-54 | Black/African-American | M | 0.78 | 0.72 | — | 0.84 | 0.00 |
| 55-64 | Black/African-American | M | 0.87 | 0.82 | — | 0.92 | 0.00 |
| 65+ | Black/African-American | M | 0.89 | 0.83 | — | 0.95 | 0.00 |
| 13-24 | Black/African-American | F | 0.90 | 0.51 | — | 1.58 | 0.71 |
| 25-34 | Black/African-American | F | 0.94 | 0.76 | — | 1.16 | 0.58 |
| 35-44 | Black/African-American | F | 0.82 | 0.72 | — | 0.94 | 0.00 |
| 45-54 | Black/African-American | F | 0.78 | 0.71 | — | 0.87 | 0.00 |
| 55-64 | Black/African-American | F | 0.83 | 0.76 | — | 0.90 | 0.00 |
| 65+ | Black/African-American | F | 0.98 | 0.89 | — | 1.08 | 0.67 |

<sup>1</sup>Change in relative mortality risk for a group is determined by dividing the relative mortality risk in 2020 by the relative mortality risk in 2018-19
<sup>2</sup>Refers to assigned sex at birth among persons with diagnosed HIV and sex determined at death. M denotes “Male,” F denotes “Female.”

**Table C6:** Mortality rates^1^ among the general Hispanic/Latino^2^ population aged ≥13 years, stratified by age and sex^3^, 2018–2020—United States

| Age | Race/Ethnicity | Sex<sup>3</sup> | Mortality Rate, 2018-19 | Mortality Rate, 2020 | Change from 2018-19 (Absolute) | Change from 2018-19 (%) | Change from 2018-19 (%), 95% confidence interval |  |
| --- | --- | --- | --- | --- | --- | --- | --- | --- |
| 13-24 | Hispanic/Latino | M | 76.2 | 98.7 | 22.5 | 29.5% | 25.4% | — 33.7% |
| 25-34 | Hispanic/Latino | M | 134.2 | 185.7 | 51.5 | 38.4% | 34.8% | — 42.1% |
| 35-44 | Hispanic/Latino | M | 178.3 | 274.0 | 95.8 | 53.7% | 50.1% | — 57.4% |
| 45-54 | Hispanic/Latino | M | 348.9 | 538.1 | 189.2 | 54.2% | 51.4% | — 57.1% |
| 55-64 | Hispanic/Latino | M | 798.2 | 1220.6 | 422.4 | 52.9% | 50.7% | — 55.2% |
| 65+ | Hispanic/Latino | M | 3,086.7 | 4373.5 | 1286.7 | 41.7% | 40.5% | — 42.9% |
| 65-74 | Hispanic/Latino | M | 1,671.1 | 2562.1 | 891.0 | 53.3% | 51.2% | — 55.5% |
| 75-84 | Hispanic/Latino | M | 3,974.0 | 5615.5 | 1641.5 | 41.3% | 39.3% | — 43.3% |
| 85+ | Hispanic/Latino | M | 10,228.5 | 13436.2 | 3207.7 | 31.4% | 29.4% | — 33.3% |
| 13-24 | Hispanic/Latino | F | 26.9 | 32.9 | 6.0 | 22.1% | 15.4% | — 29.2% |
| 25-34 | Hispanic/Latino | F | 50.5 | 67.0 | 16.5 | 32.6% | 26.7% | — 38.8% |
| 35-44 | Hispanic/Latino | F | 86.4 | 115.4 | 29.0 | 33.5% | 28.8% | — 38.5% |
| 45-54 | Hispanic/Latino | F | 182.8 | 250.1 | 67.3 | 36.8% | 33.2% | — 40.6% |
| 55-64 | Hispanic/Latino | F | 435.9 | 614.6 | 178.7 | 41.0% | 38.2% | — 43.8% |
| 65+ | Hispanic/Latino | F | 2,559.9 | 3355.7 | 795.8 | 31.1% | 30.0% | — 32.2% |
| 65-74 | Hispanic/Latino | F | 1,027.9 | 1439.9 | 412.0 | 40.1% | 37.7% | — 42.5% |
| 75-84 | Hispanic/Latino | F | 2,806.0 | 3776.7 | 970.8 | 34.6% | 32.6% | — 36.6% |
| 85+ | Hispanic/Latino | F | 9,131.8 | 11447.7 | 2315.9 | 25.4% | 23.8% | — 26.9% |

<sup>1</sup>Rates calculated by dividing deaths in a group by overall group population. All rates are per 100,000 population.
<sup>2</sup>Hispanic/Latino persons can be of any race.
<sup>3</sup>Refers to sex determined at death. M denotes “Male,” F denotes “Female.”

**Table C7:** Mortality rates^1^ among Hispanic/Latino^2^ persons with diagnosed HIV (PWDH) aged ≥13 years, stratified by age and sex^3^, 2018–2020—United States

| Age | Race/Ethnicity | Sex<sup>3</sup> | Mortality Rate, 2018-19 | Mortality Rate, 2020 | Change from 2018-19 (Absolute) | Change from 2018-19 (%) | Change from 2018-19 (%), 95% confidence interval |  |
| --- | --- | --- | --- | --- | --- | --- | --- | --- |
| 13-24 | Hispanic/Latino | M | 323.5 | 424.6 | 101.1 | 31.2% | -20.8% | — 117.5% |
| 25-34 | Hispanic/Latino | M | 509.6 | 532.8 | 23.3 | 4.6% | -12.1% | — 24.4% |
| 35-44 | Hispanic/Latino | M | 683.9 | 692.4 | 8.5 | 1.2% | -11.6% | — 15.9% |
| 45-54 | Hispanic/Latino | M | 1,029.5 | 1088.5 | 59.0 | 5.7% | -4.4% | — 17.0% |
| 55-64 | Hispanic/Latino | M | 1,768.4 | 1832.1 | 63.7 | 3.6% | -4.9% | — 12.9% |
| 65+ | Hispanic/Latino | M | 3,322.8 | 3763.7 | 440.9 | 13.3% | 2.7% | — 25.0% |
| 13-24 | Hispanic/Latino | F | 377.0 | 485.4 | 108.4 | 28.7% | -56.7% | — 283.2% |
| 25-34 | Hispanic/Latino | F | 650.9 | 841.1 | 190.1 | 29.2% | -13.3% | — 92.6% |
| 35-44 | Hispanic/Latino | F | 801.6 | 740.2 | -61.4 | -7.7% | -31.0% | — 23.5% |
| 45-54 | Hispanic/Latino | F | 1,156.8 | 1293.2 | 136.4 | 11.8% | -7.5% | — 35.2% |
| 55-64 | Hispanic/Latino | F | 1,684.8 | 1883.1 | 198.3 | 11.8% | -4.9% | — 31.4% |
| 65+ | Hispanic/Latino | F | 2,452.8 | 3297.8 | 845.0 | 34.5% | 11.9% | — 61.6% |

<sup>1</sup>Rates calculated by dividing deaths in a group by overall group population, in this case PWDH deaths divided by diagnosed HIV prevalence in the specified group. All rates are per 100,000 population.
<sup>2</sup>Hispanic/Latino persons can be of any race.
<sup>3</sup>Refers to assigned sex at birth. M denotes “Male,” F denotes “Female.”

**Table C8:** Excess mortality^1^ among Hispanic/Latino^2^ persons with diagnosed HIV (PWDH) aged ≥13 years, stratified by age and sex^3^, 2018–2020—United States

| Age | Race/ethnicity | Sex | Excess Deaths, 2020 | 95% confidence interval |  |  |
| --- | --- | --- | --- | --- | --- | --- |
| 13-24 | Hispanic/Latino | M | 6 | -4 | — | 21 |
| 25-34 | Hispanic/Latino | M | 9 | -23 | — | 46 |
| 35-44 | Hispanic/Latino | M | 4 | -36 | — | 50 |
| 45-54 | Hispanic/Latino | M | 30 | -23 | — | 90 |
| 55-64 | Hispanic/Latino | M | 28 | -38 | — | 100 |
| 65+ | Hispanic/Latino | M | 75 | 15 | — | 142 |
| 13-24 | Hispanic/Latino | F | 1 | -2 | — | 11 |
| 25-34 | Hispanic/Latino | F | 9 | -4 | — | 28 |
| 35-44 | Hispanic/Latino | F | -5 | -22 | — | 17 |
| 45-54 | Hispanic/Latino | F | 17 | -11 | — | 51 |
| 55-64 | Hispanic/Latino | F | 24 | -109 | — | 65 |
| 65+ | Hispanic/Latino | F | 51 | 18 | — | 91 |

<sup>1</sup>Excess mortality in a group is calculated by multiplying the total population size, the 2018-19 mortality rate, and the calculated percent change in the mortality rate in 2020 from 2018-19.
<sup>2</sup>Hispanic/Latino persons can be of any race.
<sup>3</sup>Refers to assigned sex at birth. M denotes “Male,” F denotes “Female.”

**Table C9:**
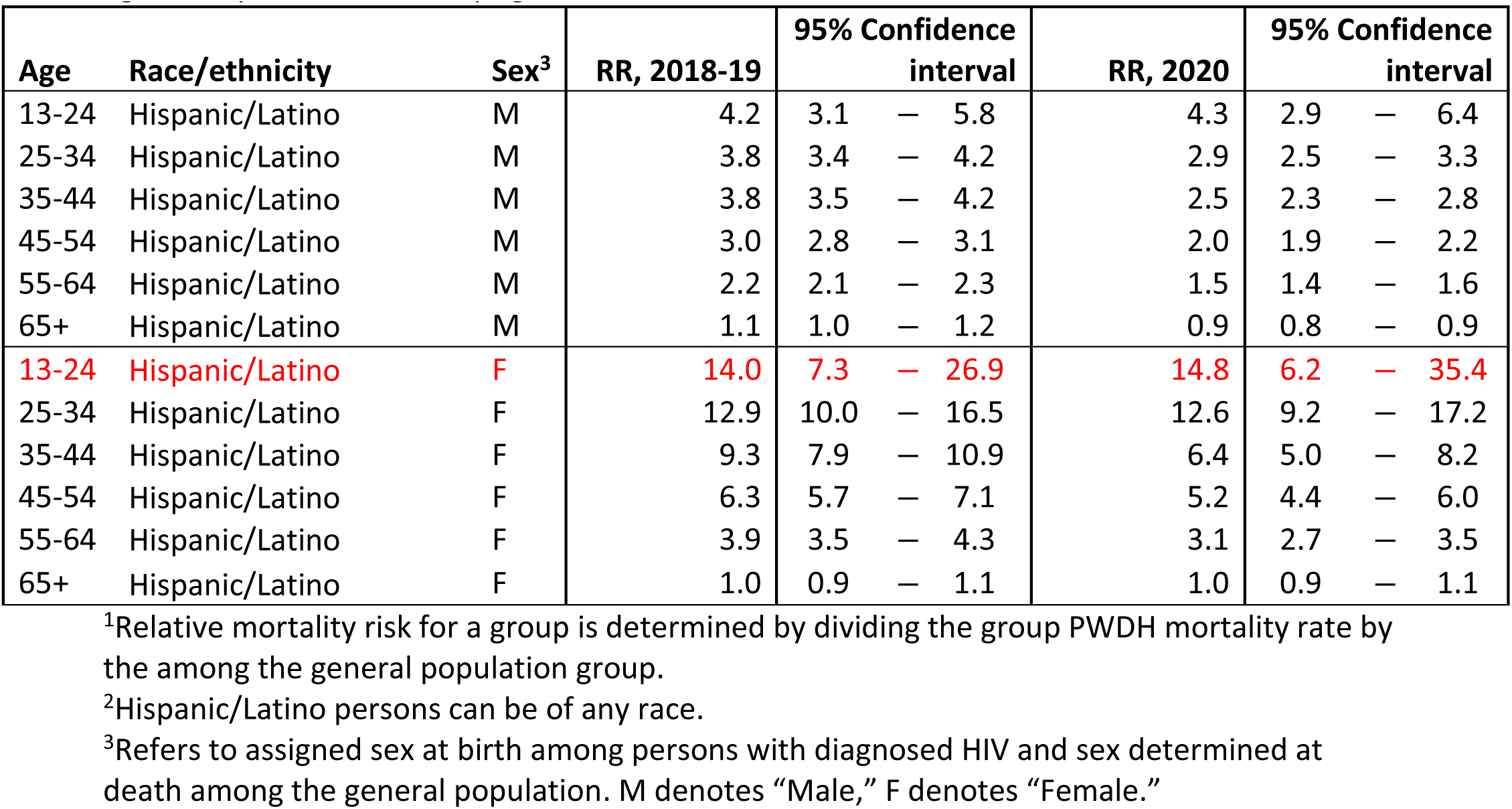
Relative mortality risk^1^ among Hispanic/Latino^2^ persons with diagnosed HIV (PWDH) aged ≥13 years, stratified by age and sex^3^, 2018–2020—United States

| Age | Race/ethnicity | Sex<sup>3</sup> | RR, 2018-19 | 95% Confidence interval |  |  | RR, 2020 | 95% Confidence interval |  |  |
| --- | --- | --- | --- | --- | --- | --- | --- | --- | --- | --- |
| 13-24 | Hispanic/Latino | M | 4.2 | 3.1 | — | 5.8 | 4.3 | 2.9 | — | 6.4 |
| 25-34 | Hispanic/Latino | M | 3.8 | 3.4 | — | 4.2 | 2.9 | 2.5 | — | 3.3 |
| 35-44 | Hispanic/Latino | M | 3.8 | 3.5 | — | 4.2 | 2.5 | 2.3 | — | 2.8 |
| 45-54 | Hispanic/Latino | M | 3.0 | 2.8 | — | 3.1 | 2.0 | 1.9 | — | 2.2 |
| 55-64 | Hispanic/Latino | M | 2.2 | 2.1 | — | 2.3 | 1.5 | 1.4 | — | 1.6 |
| 65+ | Hispanic/Latino | M | 1.1 | 1.0 | — | 1.2 | 0.9 | 0.8 | — | 0.9 |
| **13-24** | **Hispanic/Latino** | **F** | **14.0** | **7.3** | **—** | **26.9** | **14.8** | **6.2** | **—** | **35.4** |
| 25-34 | Hispanic/Latino | F | 12.9 | 10.0 | — | 16.5 | 12.6 | 9.2 | — | 17.2 |
| 35-44 | Hispanic/Latino | F | 9.3 | 7.9 | — | 10.9 | 6.4 | 5.0 | — | 8.2 |
| 45-54 | Hispanic/Latino | F | 6.3 | 5.7 | — | 7.1 | 5.2 | 4.4 | — | 6.0 |
| 55-64 | Hispanic/Latino | F | 3.9 | 3.5 | — | 4.3 | 3.1 | 2.7 | — | 3.5 |
| 65+ | Hispanic/Latino | F | 1.0 | 0.9 | — | 1.1 | 1.0 | 0.9 | — | 1.1 |

<sup>1</sup>Relative mortality risk for a group is determined by dividing the group PWDH mortality rate by the among the general population group.
<sup>2</sup>Hispanic/Latino persons can be of any race.
<sup>3</sup>Refers to assigned sex at birth among persons with diagnosed HIV and sex determined at death among the general population. M denotes “Male,” F denotes “Female.”

**Table C10:**
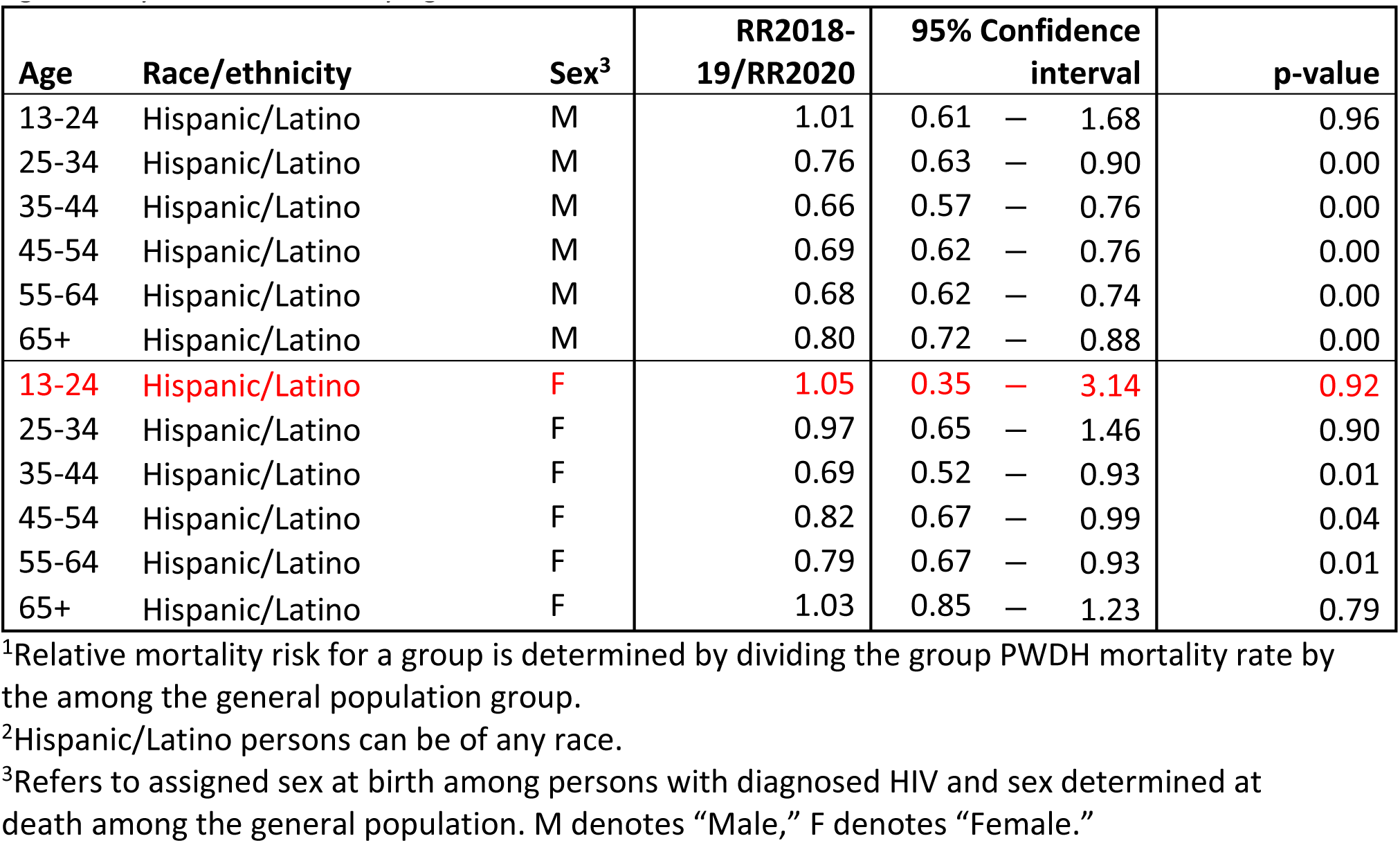
Change relative mortality risk^1^ Hispanic/Latino^2^ persons with diagnosed HIV (PWDH) aged ≥13 years, stratified by age and sex^3^, 2018–2020—United States

**Table C11:** Mortality rates^1^ among the general White population aged ≥13 years, stratified by age and sex^2^, 2018–2020—United States

| Age | Race/Ethnicity | Sex<sup>2</sup> | Mortality Rate, 2018-19 | Mortality Rate, 2020 | Change from 2018-19 (Absolute) | Change from 2018-19 (%) | Change from 2018-19 (%), 95% confidence interval |  |  |
| --- | --- | --- | --- | --- | --- | --- | --- | --- | --- |
| 13-24 | White | M | 80.8 | 89.5 | 8.7 | 10.7% | 8.3% | — | 13.2% |
| 25-34 | White | M | 183.3 | 213.6 | 30.3 | 16.5% | 14.8% | — | 18.3% |
| 35-44 | White | M | 266.7 | 319.6 | 52.9 | 19.8% | 18.3% | — | 21.4% |
| 45-54 | White | M | 505.4 | 575.6 | 70.1 | 13.9% | 12.8% | — | 14.9% |
| 55-64 | White | M | 1,116.2 | 1251.8 | 135.6 | 12.1% | 11.5% | — | 12.8% |
| 65+ | White | M | 4,433.0 | 4955.1 | 522.1 | 11.8% | 11.5% | — | 12.1% |
| 65-74 | White | M | 2,186.0 | 2455.1 | 269.1 | 12.3% | 11.8% | — | 12.8% |
| 75-84 | White | M | 5,252.7 | 5929.1 | 676.4 | 12.9% | 12.4% | — | 13.3% |
| 85+ | White | M | 15,260.4 | 17096.4 | 1836.1 | 12.0% | 11.6% | — | 12.5% |
| 13-24 | White | F | 36.3 | 37.9 | 1.5 | 4.2% | 0.7% | — | 7.9% |
| 25-34 | White | F | 87.5 | 98.7 | 11.1 | 12.7% | 10.2% | — | 15.2% |
| 35-44 | White | F | 152.1 | 175.7 | 23.6 | 15.5% | 13.6% | — | 17.5% |
| 45-54 | White | F | 316.0 | 350.2 | 34.2 | 10.8% | 9.5% | — | 12.1% |
| 55-64 | White | F | 678.5 | 748.8 | 70.3 | 10.4% | 9.6% | — | 11.2% |
| 65+ | White | F | 3,989.7 | 4371.3 | 381.5 | 9.6% | 9.3% | — | 9.8% |
| 65-74 | White | F | 1,434.9 | 1586.6 | 151.7 | 10.6% | 10.0% | — | 11.2% |
| 75-84 | White | F | 3,892.3 | 4304.2 | 411.9 | 10.6% | 10.1% | — | 11.1% |
| 85+ | White | F | 13,564.9 | 15275.6 | 1710.6 | 12.6% | 12.3% | — | 13.0% |

<sup>1</sup>Rates calculated by dividing deaths in a group by overall group population. All rates are per 100,000 population.
<sup>2</sup>Refers to sex determined at death. M denotes “Male,” F denotes “Female.”

**Table C12:** Mortality rates^1^ among White persons with diagnosed HIV (PWDH) aged ≥13 years, stratified by age and sex^2^, 2018–2020—United States

| Age | Race/Ethnicity | Sex<sup>2</sup> | Mortality Rate, 2018-19 | Mortality Rate, 2020 | Change from 2018-19 (Absolute) | Change from 2018-19 (%) | Change from 2018-19 (%), 95% confidence interval |  |  |
| --- | --- | --- | --- | --- | --- | --- | --- | --- | --- |
| 13-24 | White | M | 543.1 | 523.2 | -19.9 | -3.7% | -46.3% | — | 72.9% |
| 25-34 | White | M | 636.6 | 794.6 | 157.9 | 24.8% | 4.8% | — | 48.7% |
| 35-44 | White | M | 900.2 | 991.9 | 91.7 | 10.2% | -2.8% | — | 25.0% |
| 45-54 | White | M | 1,415.0 | 1405.1 | -9.9 | -0.7% | -8.2% | — | 7.4% |
| 55-64 | White | M | 1,866.8 | 1887.1 | 20.3 | 1.1% | -4.6% | — | 7.1% |
| 65+ | White | M | 3,329.5 | 3344.5 | 15.0 | 0.5% | -5.7% | — | 7.0% |
| 13-24 | White | F | 725.1 | 775.2 | 50.1 | 6.9% | -59.7% | — | 183.8% |
| 25-34 | White | F | 1,361.8 | 1585.8 | 224.1 | 16.5% | -13.2% | — | 56.2% |
| 35-44 | White | F | 1,527.6 | 1562.3 | 34.7 | 2.3% | -17.8% | — | 27.3% |
| 45-54 | White | F | 1,996.2 | 1762.9 | -233.2 | -11.7% | -25.3% | — | 4.4% |
| 55-64 | White | F | 2,357.5 | 2186.5 | -171.0 | -7.3% | -20.0% | — | 7.6% |
| 65+ | White | F | 3,167.5 | 3575.0 | 407.5 | 12.9% | -6.4% | — | 36.1% |

<sup>1</sup>Rates calculated by dividing deaths in a group by overall group population, in this case PWDH deaths divided by diagnosed HIV prevalence in the specified group. All rates are per 100,000 population.
<sup>2</sup>Refers to assigned sex at birth. M denotes “Male,” F denotes “Female.”

**Table C13:** Excess mortality^1^ among White persons with diagnosed HIV (PWDH) aged ≥13 years, stratified by age and sex^2^, 2018–2020—United States

| Age | Race/ethnicity | Sex<sup>2</sup> | Excess Deaths, 2020 | 95% confidence interval |  |  |
| --- | --- | --- | --- | --- | --- | --- |
| 13-24 | White | M | -1 | -8 | — | 12 |
| 25-34 | White | M | 41 | 8 | — | 79 |
| 35-44 | White | M | 35 | -10 | — | 85 |
| 45-54 | White | M | -6 | -73 | — | 66 |
| 55-64 | White | M | 19 | -79 | — | 121 |
| 65+ | White | M | 7 | -84 | — | 103 |
| 13-24 | White | F | 0 | -3 | — | 10 |
| 25-34 | White | F | 10 | -8 | — | 33 |
| 35-44 | White | F | 3 | -21 | — | 32 |
| 45-54 | White | F | -25 | -55 | — | 10 |
| 55-64 | White | F | -20 | -55 | — | 21 |
| 65+ | White | F | 20 | -10 | — | 57 |

<sup>1</sup>Excess mortality in a group is calculated by multiplying the total population size, the 2018-19 mortality rate, and the calculated percent change in the mortality rate in 2020 from 2018-19.
<sup>2</sup>Refers to assigned sex at birth. M denotes “Male,” F denotes “Female.”

**Table C14:** Relative mortality risk^1^ among White persons with diagnosed HIV (PWDH) aged ≥13 years, stratified by age and sex^2^, 2018–2020—United States

| Age | Race/ethnicity | Sex<sup>2</sup> | RR, 2018-19 | 95% Confidence interval |  |  | RR, 2020 | 95% Confidence interval |  |  |
| --- | --- | --- | --- | --- | --- | --- | --- | --- | --- | --- |
| 13-24 | White | M | 6.7 | 4.9 | — | 9.3 | 5.9 | 3.6 | — | 9.5 |
| 25-34 | White | M | 3.5 | 3.1 | — | 3.9 | 3.7 | 3.2 | — | 4.3 |
| 35-44 | White | M | 3.4 | 3.1 | — | 3.6 | 3.1 | 2.8 | — | 3.4 |
| 45-54 | White | M | 2.8 | 2.7 | — | 2.9 | 2.4 | 2.3 | — | 2.6 |
| 55-64 | White | M | 1.7 | 1.6 | — | 1.7 | 1.5 | 1.4 | — | 1.6 |
| 65+ | White | M | 0.8 | 0.7 | — | 0.8 | 0.7 | 0.6 | — | 0.7 |
| 13-24 | White | F | 20.0 | 11.4 | — | 35.1 | 20.5 | 9.2 | — | 45.5 |
| 25-34 | White | F | 15.6 | 13.0 | — | 18.6 | 16.1 | 12.7 | — | 20.3 |
| 35-44 | White | F | 10.1 | 8.8 | — | 11.4 | 8.9 | 7.4 | — | 10.6 |
| 45-54 | White | F | 6.3 | 5.8 | — | 6.9 | 5.0 | 4.4 | — | 5.8 |
| 55-64 | White | F | 3.5 | 3.2 | — | 3.8 | 2.9 | 2.6 | — | 3.3 |
| 65+ | White | F | 0.8 | 0.7 | — | 0.9 | 0.8 | 0.7 | — | 0.9 |

<sup>1</sup>Relative mortality risk for a group is determined by dividing the group PWDH mortality rate by the among the general population group.
<sup>2</sup>Refers to assigned sex at birth among persons with diagnosed HIV and sex determined at death among the general population. M denotes “Male,” F denotes “Female.”

**Table C15:** Change relative mortality risk^1^ among White persons with diagnosed HIV (PWDH) aged ≥13 years, stratified by age and sex^2^, 2018–2020—United States

| Age | Race/ethnicity | Sex<sup>2</sup> | RR2018-19/RR2020 | 95% Confidence interval |  |  | p-value |
| --- | --- | --- | --- | --- | --- | --- | --- |
| 13-24 | White | M | 0.87 | 0.48 | — | 1.56 | 0.64 |
| 25-34 | White | M | 1.07 | 0.90 | — | 1.28 | 0.44 |
| 35-44 | White | M | 0.92 | 0.81 | — | 1.04 | 0.19 |
| 45-54 | White | M | 0.87 | 0.81 | — | 0.94 | 0.00 |
| 55-64 | White | M | 0.90 | 0.85 | — | 0.96 | 0.00 |
| 65+ | White | M | 0.90 | 0.84 | — | 0.96 | 0.00 |
| 13-24 | White | F | 1.03 | 0.39 | — | 2.72 | 0.96 |
| 25-34 | White | F | 1.03 | 0.77 | — | 1.39 | 0.83 |
| 35-44 | White | F | 0.89 | 0.71 | — | 1.10 | 0.28 |
| 45-54 | White | F | 0.80 | 0.67 | — | 0.94 | 0.01 |
| 55-64 | White | F | 0.84 | 0.72 | — | 0.97 | 0.02 |
| 65+ | White | F | 1.03 | 0.85 | — | 1.24 | 0.76 |

<sup>1</sup>Relative mortality risk for a group is determined by dividing the group PWDH mortality rate by the among the general population group.
<sup>2</sup>Refers to assigned sex at birth among persons with diagnosed HIV and sex determined at death among the general population. M denotes “Male,” F denotes “Female.”

## Notes

### Competing Interest Statement

The authors have declared no competing interest.

### Funding Statement

This study was funded by the Centers for Disease Control and Prevention

